# Which Features of Medication Double-Check Procedures are Considered Desirable and Implemented in Practice? A Cross-Sectional Web-Based Vignette Study among Nurses in Japan

**DOI:** 10.64898/2026.10.04.26364685

**Authors:** Ryoi Morimoto, Tetsuro Sakura, Keisuke Sato, Kazuhiro Ueda, Rina Kagawa

**Affiliations:** Department of General Systems Studies, Graduate School of Arts and Sciences, The University of Tokyo, Tokyo, Japan; Artificial Intelligence Research Center, National Institute of Advanced Industrial Science and Technology, Ibaraki, Japan

**Keywords:** Safety, Medication Errors, Nurses

## Abstract

**Objective:** Nurses’ specific medication-administration double-check procedures vary widely; while independent double-checks (IDCs) are considered effective and recommended, they are implemented insufficiently in practice. This study aimed to classify these procedures by their component features and determine which features nurses consider desirable and perceive as implemented in clinical practice.

**Design:** Cross-sectional web-based vignette study

**Setting, participants, and methods:** Nurses working in Japanese hospitals or clinics participated in this study, for which we created 84 scenarios by combining checking procedures’ features, including independence between checkers, across the calculation, preparation, and administration stages. For each scenario, participants self-reported the desirability of the procedure and its perceived implementation frequency in their clinical setting. Linear mixed-effects models assessed associations between checking-procedure features and each rating, and McNemar’s test assessed discrepancies between desirability and implementation frequency for IDC.

**Primary outcome:** Associations between double-check procedures’ features and their desirability and implementation frequency, and the alignment between desirability and implementation frequency for IDC

**Results:** Of 534 respondents, 265 were analyzed after prespecified attention-check exclusions. Independent checking was positively associated with desirability (*b*=0.304, 95% CI [0.255, 0.353], p<0.001), perceived implementation frequency for high-risk medications (*b*=0.140, 95% CI [0.080, 0.200], p<0.001), and non-high-risk medications (*b*=0.083, 95% CI [0.022, 0.143], p=0.008). Only 22.5% of nurses perceived IDC as the most frequently implemented procedure during the calculation stage. Moreover, 23.4% rated IDC as the most desirable, but did not report it as the most frequently implemented, whereas 8.1% showed the reverse pattern (OR=2.89, 95% CI [1.69, 4.94], p<0.001).

**Conclusions:** Although independence, a key feature of IDC, was reflected in nurses’ desirability ratings and perceived implementation frequency, IDC was not the most frequently implemented checking procedure, relative to others. Implementing IDC may require not only promoting understanding of its significance but also establishing work protocols and organizational environments that enable independent checking.

## 1 Introduction

Medication errors are a major patient safety concern [1]. The medication process involves many steps and people [2], and nurses often play a major role in its final stages, including dose calculation, medication preparation, and administration to the patient [3,4]. This study focuses on the sequence of nurses’ medication-related procedures.

Double-checking is widely recommended and implemented as a strategy to prevent medication errors [5] and is viewed positively by medical professionals, including nurses [6,7]. However, evidence that double-checking reduces medication errors is insufficient [2,8]. One report found that 50.9% of cases in which double-checking was performed still failed to prevent medication errors [9]. An underlying issue is that there is no widely accepted, standardized definition of “double-checking”; different procedures, such as self-rechecking, checking jointly by two people, and independent checking by two people, are treated under the same label [2,6,7]. These procedures can be distinguished by features such as the number of people involved in checking, the number of times the same item is checked or compared, and the independence between checkers [10].

An independent double-check (IDC) is a procedure in which two checkers each compare the same target without referring to the other’s result and then compare their results afterward [11]. Learning the first checker’s result in advance can bias the second checker toward confirming that result, thereby making discrepancies less likely to be detected (i.e., confirmation bias [12]). Against this background, various guidelines explicitly identify independence between checkers as a requirement for double-checking [11,13]. Indeed, non-IDC has been reported not to reduce errors, compared with single checking [14], whereas IDC has been reported to facilitate error detection, compared with single checking [15]. Nevertheless, double-checking is rarely performed independently in clinical practice [14,16]. Even procedures intended as IDC may fail to meet IDC requirements because checkers consult prior information [17]. Nurses have also been reported to regard checking together with another person, rather than independence, as an essential feature of double-checking [7].

With the above in mind, rather than treating double-checking as a single, uniform practice, it is necessary to deconstruct checking procedures into their individual features and determine which features nurses consider desirable and which are actually implemented in clinical practice. Previous research has not sufficiently clarified which features of checking procedures are associated with perceived desirability or implementation [7]. Moreover, because double-checking requires personnel and time, it is important to target it to high-risk situations, rather than apply it uniformly to all medication administrations [11,13,18]. It is important to understand how checking procedures’ individual features are associated with their desirability and perceived implementation frequency in routine medication-administration settings, as doing so could provide a basis for determining which types of checks should be prioritized in which situations, so that double-checking functions more effectively under limited personnel and time resources.

It is also necessary to clarify why IDC is insufficiently implemented. As a first step, the relationship between IDC’s perceived desirability and implementation frequency should be examined. However, whether these correspond within the same population has not been established, so it remains unclear whether insufficient implementation of IDC reflects a failure to regard IDC as a desirable procedure, or whether nurses see it as desirable despite perceiving it as insufficiently implemented. Distinguishing these possibilities provides a basis for understanding why IDC is insufficiently implemented in practice, for considering whether implementation requires a greater understanding of its significance, or if changes to clinical procedures and organizational environments are needed to enhance feasibility.

Based on a sample of nurses in Japan, this study organized medication-related checking procedures according to three features̶independence, number of checkers, and number of checks—and examined which are considered desirable and which are perceived as being implemented in clinical settings. It also considered whether desirability and perceived implementation align for IDC. Because the direct observation of actual implementation frequency is not straightforward, we assessed how frequently nurses perceived each checking procedure being implemented in their clinical setting.

Accordingly, this study addressed the following two research questions:

1. How are independence, the number of checkers, and number of checks in medication-related checking procedures associated with desirability for high-risk medication administration and with implementation frequency for high- and non-high-risk medication administration?
2. For IDC, do ratings of desirability and perceptions of implementation frequency align?

## 2 Methods

### 2.1 Study design and participants

This study was a cross-sectional web-based vignette survey of nurses in Japan. Participants were presented with scenarios describing medication-administration checking procedures and rated both the desirability of each checking procedure and its perceived implementation frequency in their own clinical setting. Vignettes describing specific situations have been used in previous studies of double-checking [7] and, compared with directly asking about abstract concepts, can reduce variation in the situations respondents imagine when making judgments [19,20].

Participants were nurses working in hospitals, clinics, or similar healthcare facilities in Japan. To obtain responses based on a minimum level of practical experience, continuous employment at a hospital or clinic during the year preceding the survey was an inclusion criterion. Recruitment was conducted through a commercial online survey panel in Japan. Recruitment and data collection were conducted from October to November 2025.The survey company distributed invitations to panel members registered either as nurses or in the broader occupational category of healthcare-related professionals. Participants who completed the survey received points as an incentive according to the survey company’s standard rules. The survey company requested that its identity not be disclosed when the numbers of individuals in each stage of recruitment; therefore, the company is not named in this article.

Because this study was exploratory and aimed to examine associations among features of checking procedures and their evaluations, we prioritized obtaining a sufficient number of responses from nurses with different levels of clinical experience and targeted a sample size of 300. Five categories of clinical experience were defined (0–4, 5–9, 10–14, 15–19, and ≥20 years). Recruitment was conducted so that the categories were represented approximately equally and, to the extent possible, sex imbalance was minimized.

Appendix A provides an English translation of the original Japanese study plan that was submitted for ethical review before data collection.

### 2.2 Structuring medication-related checking procedures and developing the scenarios

Based on Dickinson et al. [16], medication-related procedures performed by nurses were structured into three stages: calculation, preparation, and administration. For each stage, checking procedures that may be used in practice were systematically characterized in terms of three features: the number of people involved in checking, the number of checks, and independence between checkers. With reference to the definition of IDC provided by Australian Nursing and Midwifery Federation (Victorian Branch) [11], checking was classified as “independent” when two checkers completed checks without sharing their results; however, independence defined as two checkers each checking the same target without referring to each other’s judgment results could be explicitly specified only for the calculation stage. This resulted in seven levels for the calculation stage, four for the preparation stage, and three for the administration stage, yielding 7 × 4 × 3=84 medication-administration scenarios (Figure 1). Details of each checking format are provided in Appendix B, and all scenarios are shown in Appendix C.

**Figure 1.**
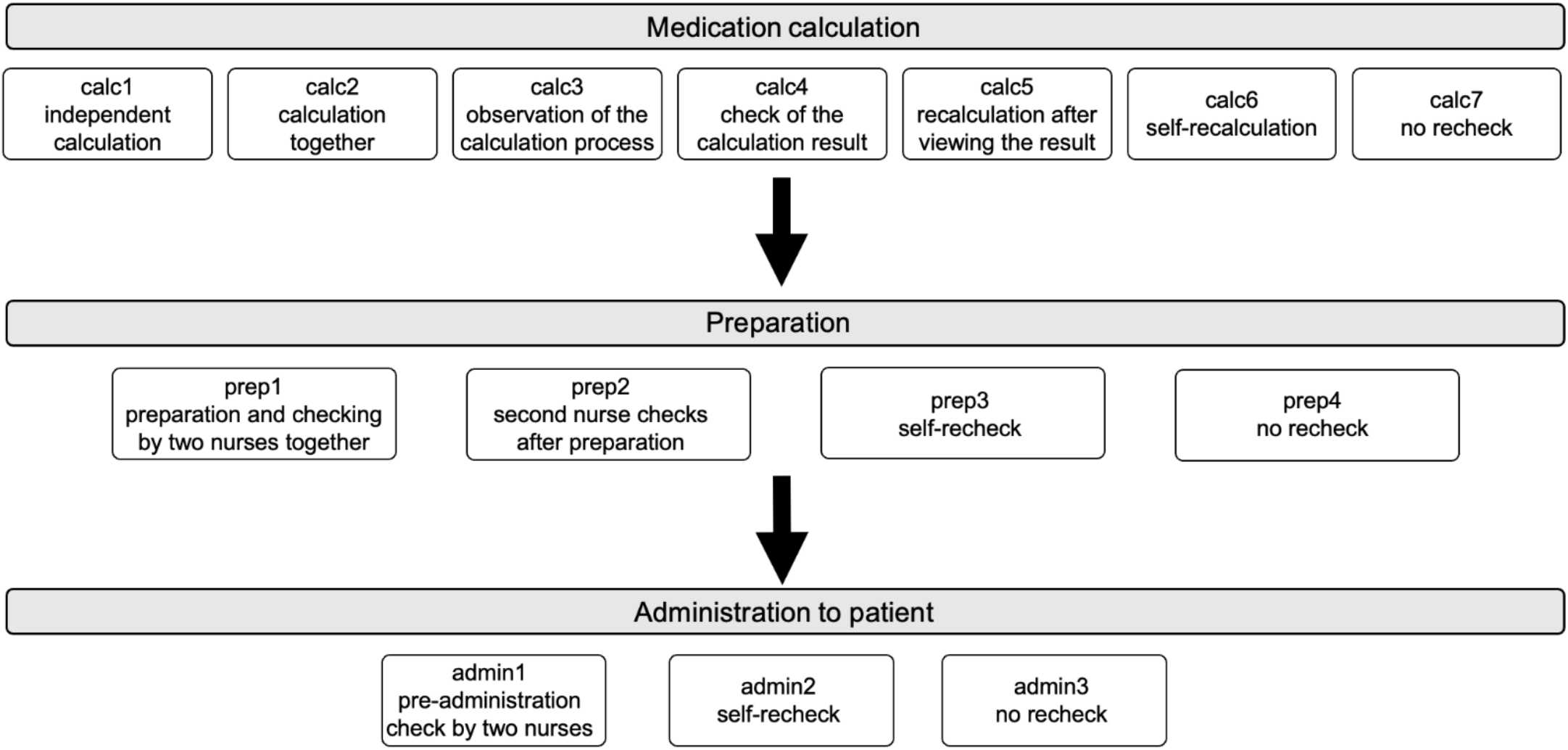
Medication-administration scenario model constructed in this study

An example of a scenario presented to participants is shown in Figure 2. In the background information common to all scenarios, a patient whose treatment required the consideration of renal function was described as receiving a fictitious high-risk medication called Karicept intravenously. Two characters, Nurses 1 and 2, were specified; in conditions involving only one checker, only Nurse 1 was involved in that stage. Each scenario presented background information followed sequentially by the procedures for the calculation, preparation, and administration stages. To ensure that participants could evaluate the differences in the checking procedures themselves, patient- and medication-related conditions were held constant across scenarios, and the medication name, dose, route, purpose, and time; calculation result; infusion-pump settings; and information provided on the medication label were explicitly stated to be correct. Basic checks corresponding to the so-called “5Rs” [21]—right medication, route, time, dose, and patient—were assumed to be performed appropriately according to each facility’s standard procedures. With these conditions held constant, only the checking formats for the calculation, preparation, and administration stages were recombined across scenarios. Prior to participants’ involvement, three nurses recruited from outside the research team, each with at least eight years of clinical experience, confirmed that the checking procedure described in each scenario could be reasonably imagined as a clinical situation.

**Figure 2:**
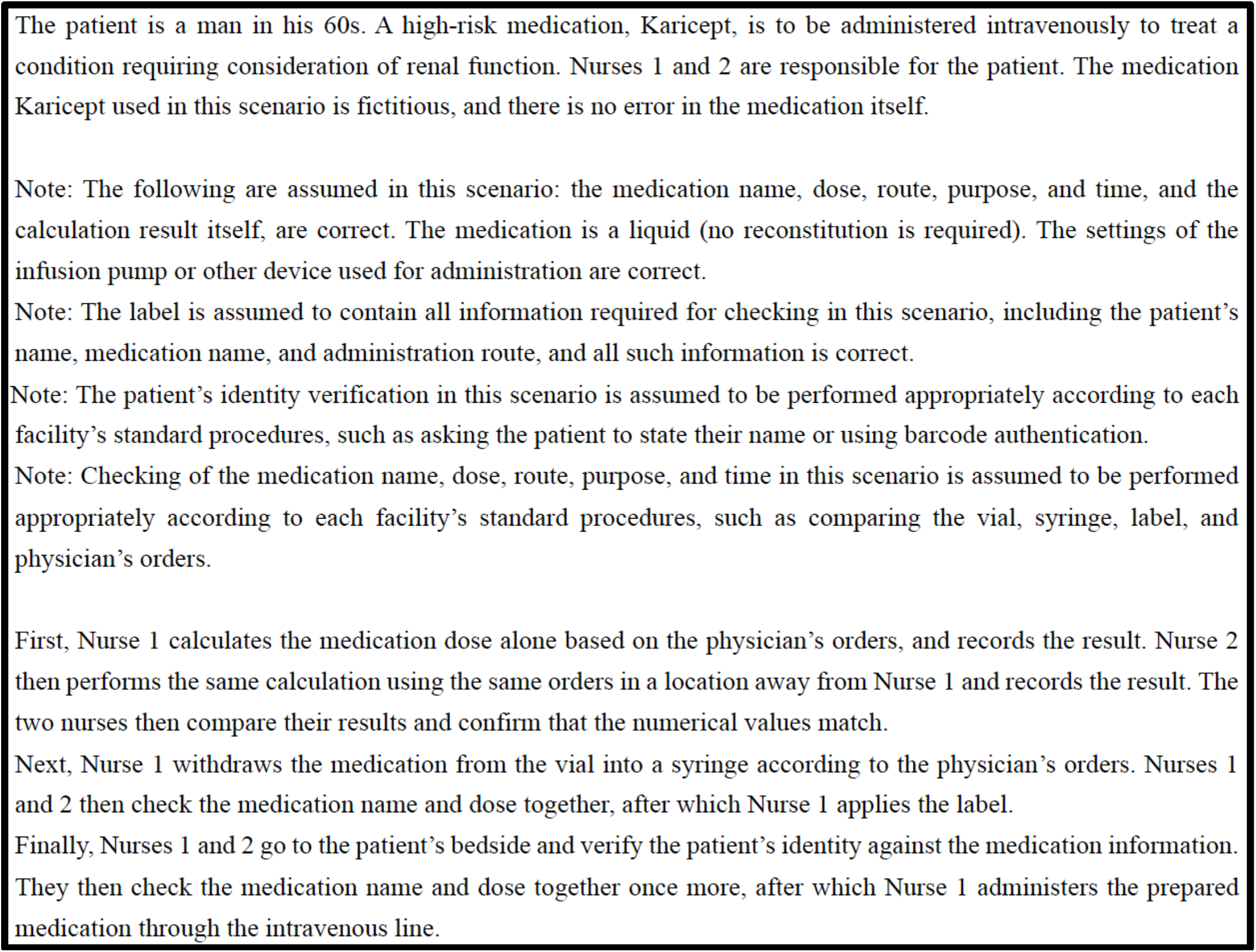
Example scenario comprising calc1 (independent calculation), prep1 (preparation and checking by two nurses together), and admin1 (pre-administration check by two nurses) (see Appendix B).

### 2.3 Scenario rating items and procedure

For each scenario, participants rated the desirability of the checking procedure and how frequently they perceived it to be implemented in their clinical setting. Implementation frequency was assessed via self-reporting, rather than by direct observation of medication-administration practices. Desirability was evaluated for high-risk medication administration, where the use of double-checking warrants particular consideration. Implementation frequency was evaluated for both high- and non-high-risk medications, to capture routine practices involving non-high-risk medications.

Specifically, for desirability, participants were asked about the checking procedure presented for high-risk medication administration: “How appropriate do you think the checking method described in this scenario is?” (Q1). For implementation frequency, participants were asked about the same checking procedure for high-risk medication administration: “When administering a high-risk medication, how often is the checking method described in this scenario implemented in practice in your clinical setting?” (Q2). They were also asked to imagine using the same checking procedure when administering a non-high-risk medication: “When administering a medication other than a high-risk medication, how often is the checking method described in this scenario implemented in practice in your clinical setting?” (Q3). Q1–Q3 were answered using 7-point scales. For Q1, 1 indicated “not at all appropriate,” 4 indicated “neither appropriate nor inappropriate,” and 7 indicated “very appropriate.” For Q2 and Q3, 1 indicated “not implemented at all,” 4 indicated “a neutral midpoint,” and 7 indicated “implemented very frequently.” For Q2 and Q3, even if continuous infusions were not performed in the respondent’s clinical setting, they were instructed to imagine, respectively, high-or non-high-risk medication infusion/injection situations. If high-risk medications were not handled at all at the respondent’s clinical setting, they could select “do not administer high-risk medications” for Q2; if infusions or injections were not performed at all in the respondent’s clinical setting, they could select “do not perform infusions or injections” for Q3. If either response was selected only once, it was considered potentially accidental; for respondents who selected it two or more times, responses to the applicable question were treated as missing in the analysis.

The survey comprised three sections, which corresponded to Q1, Q2, and Q3, and the 84 scenarios were presented in a random order within each section. The survey therefore included 252 scenario-rating items in total. To detect inattentive responses, three attention-check items per section (nine in total) instructed respondents within the question text to select a specified response option [22]. Respondents who failed to select the specified option on two or more of the nine items were excluded from the analysis.

The following participant characteristics were collected: age, sex, years of nursing experience, number of beds in their clinical setting, and frequency of high- and non-high-risk medication administration. All questions and scenarios used in the study were presented in Japanese; they are presented in English in this article to facilitate readers’ understanding.

### 2.4 Statistical analyses

To examine associations between checking-procedure features and ratings, linear mixed-effects models were fitted separately with Q1 (desirability), Q2 (implementation frequency for high-risk medications), and Q3 (implementation frequency for non-high-risk medications) as the outcome variables. To account for the repeated-measures structure in which the same respondent evaluated multiple scenarios, a random intercept was included for each respondent. Three models were specified for each of Q1–Q3. Model 1 included, as fixed effects, independence in the calculation stage; the number of checkers; and the number of checks in the calculation, preparation, and administration stages. Model 2 added respondents’ characteristics to Model 1: number of beds in the clinical setting, years of nursing experience, frequency of non-high-risk medication administration, and frequency of high-risk medication administration. Model 3 added interactions between independence and each respondent’s characteristics to Model 2, to examine whether the association with independence differed by respondent characteristics. Model fit was compared using the Akaike information criterion (AIC) and Bayesian information criterion (BIC). Details of the models and definitions of the variables are provided in Appendix D.1.

To examine within-respondent alignment between desirability and implementation frequency, Q1 and Q2 were compared for the same high-risk medication-administration situation. Because the checking format with independence (IDC) was specified only in the calculation stage, this analysis focused on calc1: independent calculation (see Appendices B.2 and B.3), which corresponded to IDC in the calculation stage. Among respondents included in the analysis of perceived implementation frequency for high-risk medication administration (Q2), Q1 and Q2 responses from the same respondent were paired. For each respondent, and each of the seven calculation-stage checking formats, mean scenario ratings were calculated separately for Q1 and Q2. Each mean rating was based on ratings from the 12 scenarios combining the relevant calculation condition with the four preparation conditions and three administration conditions. For Q1 and Q2 separately, we determined whether the mean rating of scenarios containing IDC, that is, calc1 (independent calculation), ranked highest among the seven calculation conditions. Respondents were then classified into four groups according to the combination of these two determinations. Among respondents whose classifications differed between Q1 and Q2, the direction of discordance was examined using McNemar’s test.

Tests of fixed effects in the linear mixed-effects models and McNemar’s test were two-sided, with a significance level of 5%. Statistical analyses were performed using R version 4.5.2.

### 2.5 Patient and public involvement

Neither patients nor the public were involved in the design, conduct, reporting, or dissemination plans of this research.

### 2.6 Ethical considerations

This study involved human participants and was approved by The Ethical Review Committee for Experimental Research Involving Human Subjects at the Graduate School of Arts and Sciences, The University of Tokyo, Japan (approval number 1075). Participants gave their informed consent to participate in the study before taking part.

## 3 Results

### 3.1 Participants’ characteristics

The survey company distributed invitations to 27,513 panel members in the recruitment categories described above. Of these, 1,009 accessed the survey and 534 completed it. Among these 534 respondents, 269 (50.4%) failed to select the specified response option on two or more of the nine attention-check items and were excluded, leaving 265 respondents for analysis (mean age 41.55 years, SD 8.74; 55 men, 210 women; no participants selected “other” or “prefer not to say” for sex) (Table 1). For Q2, 43 respondents who selected “do not administer high-risk medications” in multiple scenarios were excluded, leaving 222 for analysis. For Q3, 10 respondents who selected “do not perform infusions or injections” in multiple scenarios were excluded, leaving 255 for analysis. In addition, 11 respondents for Q2 and 15 respondents for Q3 selected these options in only one scenario. These single selections were considered potentially accidental, and the corresponding scenario-level responses were treated as missing and omitted from the analyses.

**Table 1.** Participants’ characteristics (N=265)

|  |  | n | % |  |  | n | % |
| --- | --- | --- | --- | --- | --- | --- | --- |
| Nursing experience | <5 years | 13 | 4.9 | Medication administration frequency | Never | 7 | 2.6 |
|  | 5 to <10 years | 49 | 18.5 |  | Rarely (<1/month) | 21 | 7.9 |
|  | 10 to <15 years | 59 | 22.3 |  | Several times/month (<1/week) | 31 | 11.7 |
|  | 15 to <20 years | 64 | 24.2 |  | Several times/week (about 1 to several/week) | 64 | 24.2 |
|  | ≥20 years | 80 | 30.2 |  | Almost daily (on nearly all working days) | 142 | 53.6 |
| **Total** |  | **265** | **100.0** | **Total** |  | **265** | **100.0** |
| Beds in the clinical setting | No inpatient beds (0 beds) | 37 | 14.0 | High-risk medication frequency | Never | 47 | 17.7 |
|  | 1-19 beds | 7 | 2.6 |  | Rarely (<1/month) | 61 | 23.0 |
|  | 20-99 beds | 23 | 8.7 |  | Several times/month (<1/week) | 48 | 18.1 |
|  | 100-199 beds | 42 | 15.8 |  | Several times/week (about 1 to several/week) | 62 | 23.4 |
|  | 200-299 beds | 46 | 17.4 |  | Almost daily (on nearly all working days) | 47 | 17.7 |
|  | 300-499 beds | 58 | 21.9 |  |  |  |  |
|  | ≥500 beds | 50 | 18.9 |  |  |  |  |
|  | Other/unknown | 2 | 0.8 |  |  |  |  |
| **Total** |  | **265** | **100.0** | **Total** |  | **265** | **100.0** |

### 3.2 Factors associated with desirability and perceived implementation frequency of checking procedures

To examine how checking-procedure features and respondents’ characteristics were associated with the desirability and implementation frequency of checking procedures, linear mixed-effects models were estimated separately with Q1 (desirability), Q2 (implementation frequency for high-risk medications), and Q3 (implementation frequency for non-high-risk medications) as outcomes. Model 1 included checking-procedure features (independence, number of checkers, and number of checks), Model 2 added respondents’ characteristics, and Model 3 added further interactions between independence and respondents’ characteristics. Supplementary analyses of rating patterns by individual checking procedure are provided in Appendix D.2.

#### 3.2.1 Model construction and comparison

For all three outcomes (Q1‒Q3), AIC was lowest for Model 3, which included interaction terms, whereas BIC was lowest for Model 1. Accordingly, the basic associations between checking-procedure features and ratings are presented below based on Model 1, and the possibility that these associations differ according to respondents’ characteristics is presented based on Model 3. Full estimates for all models, including Model 2, for Q1‒Q3 are provided in Appendix D.3.

#### 3.2.2 Associations between checking-procedure features and ratings: Model 1

Independence was positively associated with desirability (*b*=0.304, 95% CI [0.255, 0.353], p<0.001), implementation frequency for high-risk medications (*b*=0.140, 95% CI [0.080, 0.200], p<0.001), and implementation frequency for non-high-risk medications (*b*=0.083, 95% CI [0.022, 0.143], p=0.008), with the coefficient decreasing in that order. In other words, independent checking was rated as more desirable than non-independent checking and perceived as being implemented more frequently for both high- and non-high-risk medications. However, the magnitude of the association with independence was largest for desirability, followed by implementation frequency for high-risk medications and then for non-high-risk medications.

For the number of checkers, checking by two people was rated as more desirable than checking by one person in all three stages (calculation: *b*=1.150, 95% CI [1.101, 1.199]; preparation: *b*=1.044, 95% CI [1.003, 1.084]; administration: *b*=0.766, 95% CI [0.726, 0.807]; all p<0.001). For implementation frequency of high-risk medications, checking by two people was perceived as more frequent than checking by one person in all stages (calculation: *b*=0.889, 95% CI [0.828, 0.949]; preparation: *b*=0.903, 95% CI [0.853, 0.953]; administration: *b*=0.244, 95% CI [0.194, 0.294]; all p<0.001). For implementation frequency of non-high-risk medications, two-person checking was associated with higher implementation frequency in the calculation (*b*=0.556, 95% CI [0.495, 0.616], p<0.001) and preparation stages (*b*=0.653, 95% CI [0.603, 0.704], p<0.001), but with lower implementation frequency in the administration stage (*b*=-0.276, 95% CI [-0.326, -0.226], p<0.001).

For the number of checks, conditions with more checks were rated as more desirable in the calculation (*b*=0.134, 95% CI [0.071, 0.196], p<0.001) and administration stages (*b*=0.067, 95% CI [0.026, 0.108], p=0.001). For implementation frequency of high-risk medications, procedures involving more checks were perceived to be implemented more frequently only in the calculation stage (*b*=0.081, 95% CI [0.004, 0.157], p=0.038). For implementation frequency of non-high-risk medications, no significant association with the number of checks was observed in any stage.

#### 3.2.3 Interactions between independence and respondent characteristics: Model 3

The interaction between years of nursing experience and independence was negative for desirability (*b*=-0.084, 95% CI [-0.123, -0.044], p<0.001), implementation frequency for high-risk medications (*b*=-0.086, 95% CI [-0.134, -0.038], p<0.001), and implementation frequency for non-high-risk medications (*b*=-0.078, 95% CI [-0.126, -0.030], p=0.001). Thus, respondents with more years of nursing experience showed smaller differences in desirability and implementation frequency between independent and non-independent checking.

The interaction between number of beds and independence was not statistically significant for desirability (*b*=-0.023, 95% CI [-0.050, 0.004], p=0.090). In contrast, positive interactions were observed for implementation frequency for both high-risk (*b*=0.084, 95% CI [0.048, 0.119], p<0.001) and non-high-risk medications (*b*=0.108, 95% CI [0.075, 0.141], p<0.001). Thus, whereas the desirability gap between independent and non-independent checking did not clearly differ by number of beds, respondents working in clinical settings with more beds showed larger differences in implementation frequency between independent and non-independent checking.

For medication-administration frequency, no significant interactions with independence were observed for either desirability or perceived implementation frequency for either high- or non-high-risk medication administration.

### 3.3 Alignment between desirability and implementation frequency for IDC

For scenarios containing IDC in the calculation stage, we examined the alignment between desirability and implementation frequency for high-risk medications (n=222). For desirability and implementation frequency separately, respondents were classified according to whether the mean rating of scenarios containing IDC ranked highest among the seven calculation conditions, and the distribution across the four resulting groups was examined.

As shown in Figure 3, many respondents (54.1%) did not rank scenarios containing IDC highest for either desirability or implementation frequency, whereas 14.4% ranked IDC highest for both. For implementation frequency, 77.5% of respondents did not rank IDC highest.

**Figure 3.**
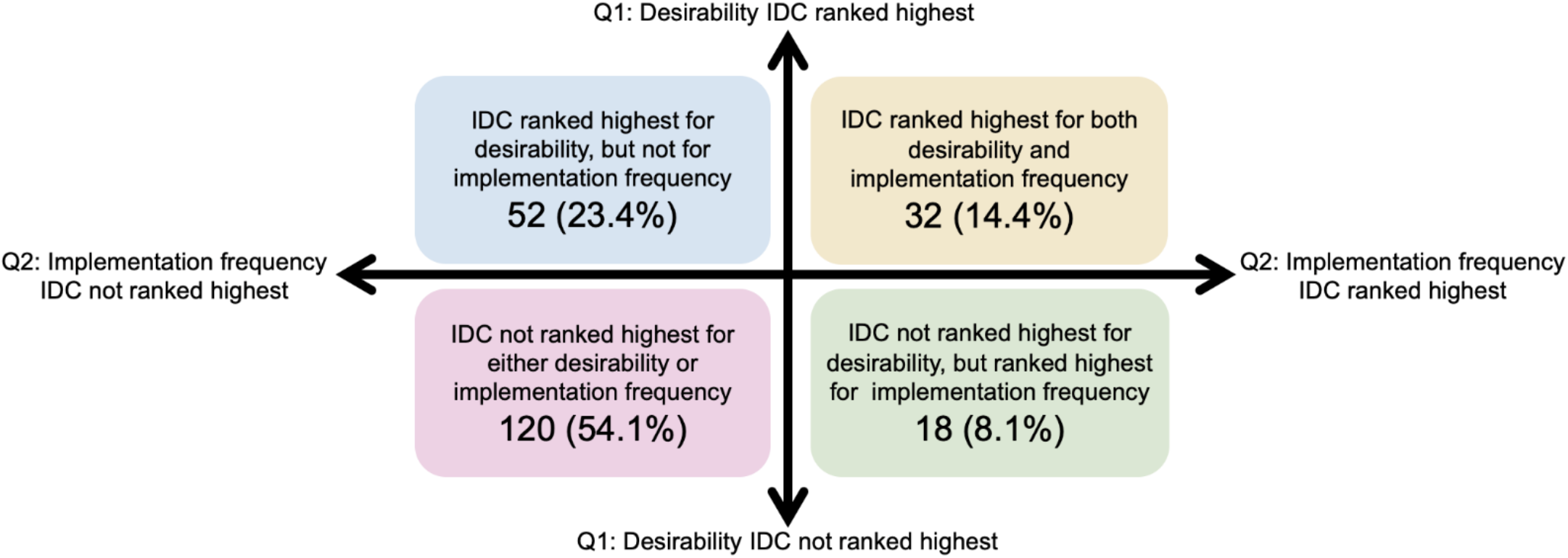
Alignment between desirability (Q1) and implementation frequency (Q2)

Among respondents whose classifications differed between the two measures, those for whom IDC ranked highest for desirability but not for implementation frequency (23.4%) outnumbered those with the reverse pattern (8.1%). McNemar’s test showed a significant directional imbalance in this discordance (*χ*^2^(1)=15.56, p<0.001), with an odds ratio based on discordant pairs of 2.89 (95% CI [1.69, 4.94]). Thus, when desirability and implementation frequency did not correspond, the more common pattern was for checking procedures containing IDC to be rated as the most desirable relative to other procedures, yet not perceived as the most frequently implemented in the clinical setting.

## 4 Discussion

In this study, we systematically represented medication-related checking procedures performed by nurses according to three features: independence, number of checkers, and number of checks. We assessed both the desirability and perceived implementation frequency for the same checking procedures. Independent checking was rated as more desirable than non-independent checking and perceived as being implemented more frequently in respondents’ clinical settings. However, 23.4% of respondents rated IDC as the most desirable checking procedure, yet did not report it as the most frequently implemented in their clinical setting.

One strength of this study is that, rather than treating double-checking as a single uniform practice, we deconstructed checking procedures into their component features and systematically compared them using 84 scenarios covering the calculation, preparation, and administration stages. Desirability and perceived implementation frequency were assessed for the same procedures while patient- and medication-related conditions were held constant. Limitations include the use of self-reported data, rather than directly observed implementation frequency; the restriction of explicit manipulation of independence to the calculation stage; and the use of relative rankings in the IDC correspondence analysis. Because this was a cross-sectional study, causal relationships cannot be inferred from associations with respondents’ characteristics. The exclusion rate for inattentive responses was 50.4%; although attention-check failure rates vary substantially across surveys [23]. The requirement to complete up to 252 scenario ratings may have contributed to inattentive responding [24]. Because these exclusions may have selected a more attentive sample, caution is warranted when generalizing the findings.

Our findings extend previous evidence on medication double-checking. Consistent with guidelines defining independence as a requirement for IDC [11,13], independent checking was rated as more desirable than non-independent checking. Beyond independence, our study also showed that checking features were associated differently with perceived implementation depending on medication risk: for non-high-risk medications, two-person checking was associated with lower perceived implementation frequency at the administration stage, while the number of checks was not associated with perceived implementation frequency at any stage. These patterns may reflect selective use of more resource-intensive checking in higher-risk situations [11,13,18]. Furthermore, although previous studies have reported limited implementation of IDC [14,16,17], we assessed desirability and perceived implementation frequency for the same procedures in the same respondents and showed that some respondents rated IDC as the most desirable procedure without reporting it as the most frequently implemented. This suggests that a limited implementation of IDC may not be explained solely by whether nurses consider it desirable.

For clinicians and policymakers involved in patient safety, these findings suggest that promoting and implementing IDC may require not only education on the significance of independence but also the establishment of workflows and organizational environments that enable independent checking. In particular, the finding of an association with independence that varies by years of nursing experience for both desirability and perceived implementation frequency, and by facility size for perceived implementation frequency, suggests that IDC’s implementation may be influenced by both individual evaluations and existing checking practices and institutional systems. Given the previous finding that even procedures intended as IDC may, in practice, fail to satisfy the requirement for independence [17], it is important to consider, at the facility level, education and work protocols that make independent checking feasible.

Future studies should determine through interviews or other methods how frequently IDC is actually implemented in clinical settings and identify specific factors that make it difficult to maintain independence. Intervention studies that manipulate the timing of results-sharing between checkers or work allocation are also needed, to examine how procedures explicitly requiring independence affect their implementation and error detection. Generalizability to other stages, countries, and regions should also be examined.

## 5 Conclusion

By deconstructing medication-administration checking procedures into their component features, this study found that nurses’ independent checks are positively associated with both desirability and perceived implementation frequency. However, only 22.5% of nurses perceived IDC as the most frequently implemented procedure in the calculation stage, and 23.4% rated IDC as the most desirable, despite not reporting it as the most frequently implemented approach in their workplaces. Thus, although independence, a key feature of IDC, is reflected in nurses’ evaluations and perceptions of implementation, it is not considered the most frequently implemented checking procedure. This suggests that translating IDC into practice may require work protocols and organizational environments that enable independent checking.

## Data Availability

All data produced in the present study are available upon reasonable request to the authors.

## STRENGTHS AND LIMITATIONS OF THIS STUDY

- Medication-administration checking procedures were systematically organized for the calculation, preparation, and administration stages, in terms of the number of checkers, number of checks, and independence between checkers, and compared using 84 vignettes.
- For the same checking procedure, desirability for high-risk medication administration and implementation frequency in the clinical setting were assessed in a paired manner; implementation frequency was also assessed for non-high-risk medications.
- Linear mixed-effects models were used to analyze associations between checking-procedure features and each rating while accounting for the repeated-measures structure in which each participant evaluated many scenarios.
- Implementation frequency was not directly observed in actual clinical settings; rather, nurses self-reported their perceptions of how frequently each procedure was implemented in their clinical setting.
- Independence between checkers could be explicitly manipulated only at the calculation stage, limiting the direct generalization of findings on independence to the preparation and administration stages.

## Contributors

RM: Conceptualization, Methodology, Investigation, Data curation, Formal analysis, Visualization, Writing – original draft.

TS: Methodology, Formal analysis, Supervision, Writing – review & editing.

KS: Software, Formal analysis, Supervision, Writing - review & editing.

KU: Conceptualization, Methodology, Supervision, Writing – review & editing, Funding acquisition.

RK: Conceptualization, Methodology, Supervision, Visualization, Writing – review & editing, Funding acquisition.

RK is the guarantor and corresponding author.

## Funding

This study was supported by JST PRESTO, Japan (JPMJPR23I3), and JSPS KAKENHI, Japan (23K16249;26K02623) for RK, and JSPS KAKENHI, Japan (25H01131) for KU. The funders had no role in the study design; data collection, analysis, or interpretation; preparation of the manuscript; or the decision to submit the manuscript for publication.

## Appendix A Original Protocol

Note: The name of the survey company has been redacted from this English translation because the company requested that it not be identified when the numbers of individuals at each stage of recruitment is reported in this manuscript.

### 1. Study title

Survey of Nurses’ Double-Check Practices and Perceptions for the Prevention of Medical Errors

### 2. Principal investigator and research team

Principal investigator: K. Ueda, Professor, Department of General Systems Studies, Graduate School of Arts and Sciences, The University of Tokyo.

Research team: R. Morimoto, second-year Master’s student, Department of General Systems Studies, Graduate School of Arts and Sciences, The University of Tokyo; R. Kagawa, Senior Research Scientist, Artificial Intelligence Research Center, National Institute of Advanced Industrial Science and Technology (AIST).

Roles: Study planning, all investigators; study implementation, all investigators; data analysis, research team members; verification and discussion of analytical results, all investigators; manuscript preparation, all investigators.

### 3. Study period and setting

Study period: From approval by the ethics review committee through 31 March 2027.

Setting: The survey will be conducted online. Approximately 300 adult nurses residing in Japan and currently working as nurses will be recruited through web platforms operated by [survey company name redacted] or similar services, and the study will be conducted online.

### 4. Study content

#### 4.1 Purpose and expected contribution

Double-checking is widely used in clinical settings as a means of preventing medication administration errors, but its implementation in practice and effectiveness remain debated. In theory, double-checking can substantially increase error-detection rates (Campbell et al., 1998). For this reason, an “independent double-check,” in which two workers perform checks independently, is regarded as an appropriate method and is recommended in guidelines (Alberta Health Services, 2019). However, in clinical practice, a “two-person simultaneous bidirectional” approach, in which two people check together verbally, is often used (Japan Council for Quality Health Care, 2023), and such approaches have been noted to involve cognitive biases and may therefore not necessarily achieve high error-detection performance (Westbrook et al., 2021).

There also appears to be uncertainty regarding which form nurses themselves regard as “correct.” For example, Schwappach et al. (2016) reported that more than half of surveyed nurses regarded “two people checking together” as an essential feature of double-checking, suggesting that the importance of independence may not be sufficiently shared.

In this study, nurses will be presented with multiple medication-administration scenarios and asked to provide subjective ratings of the “appropriateness” of each checking method and its “frequency of implementation in clinical practice.” Through these ratings, we aim to clarify which forms of double-checking nurses regard as ideal and how they perceive these forms to be implemented in actual practice. By making visible any discrepancies between perceived current practice and the ideal, the study is expected to provide insights relevant to medical-safety education and institutional interventions.

#### 4.2 Study overview

The purpose of this study is to clarify discrepancies between nurses’ perceptions of double-checking during medication administration and its perceived implementation in practice. Quantitative data will be collected through a web-based survey and analyzed statistically. Specifically, participants will be shown multiple scenarios reproducing checking behaviors used during medication administration in clinical practice. For each scenario, participants will use a seven-point scale to rate (1) whether the checking method is appropriate and (2) how frequently they perceive that checking method to be performed in clinical practice.

A scenario-based question format has also been used in previous research on double-checking (Schwappach et al., 2016). By presenting questions based on concrete situations rather than abstract terms or concepts, healthcare professionals may be better able to recall actual work experiences and provide more natural judgments and perceptions. In the present study, multiple double-check procedures observed in clinical practice will similarly be specified as concrete scenarios to elicit ratings that reflect participants’ internal judgment criteria and experience.

The responses will be examined not only by comparing mean ratings across scenarios but also by analyzing the component features of each checking method (e.g., independence, repetition, and simultaneity) to assess how these features influence ratings.

### 5. Methods

#### 5.1 Survey procedures

A web-based questionnaire will be administered to licensed nurses recruited as survey monitors through [survey company name redacted] or similar web platforms. The questionnaire will collect basic information, including age, sex, years of nursing experience, current principal clinical department, and main work setting (e.g., inpatient ward, outpatient department, operating room). Participants will then answer questions on the appropriateness and perceived implementation frequency of the checking method described in each of 84 scenarios, using seven-point rating scales.

The questionnaire consists of: (1) Basic information; (2) Questions regarding appropriateness; and (3) Questions regarding implementation frequency. The order in which the scenario texts are presented in sections (2) and (3) will be randomized for each participant.

#### 5.2 Scenarios to be evaluated

The scenarios divide the medication-administration process into the following three stages and focus on differences in checking procedures at each stage:

1. Medication-dose calculation
2. Preparation of injectable medication
3. Administration to the patient

Multiple procedural patterns are defined for each stage (7 calculation patterns × 4 preparation patterns × 3 administration patterns). Combining these patterns yields 84 scenarios (7 × 4 × 3=84). All scenarios are written as brief descriptions of medication-administration situations. For each scenario, participants will provide ratings for the appropriateness and implementation-frequency questions described above.

#### 5.3 Other data to be collected

Response time for each question.

The web-based survey will be anonymous. Before participation, participants will be informed that the expected survey duration is approximately 90 minutes and that they may pause and resume freely. The estimated duration is approximately 5 minutes for the consent form, 1 minute for basic information, and an average of 30 seconds for each of the two ratings for 84 scenarios, for a total of approximately 90 minutes. Participants will proceed at their own pace and will be instructed that they may take breaks as needed.

### 6. Study participants

Approximately 300 adult licensed nurses residing in Japan will be recruited. Because experience with and perceptions of double-checking may differ according to years of clinical experience, clinical department, and work setting (e.g., ward, outpatient department, operating room), recruitment will aim to include participants with varied backgrounds.

Years of nursing experience will be divided into five categories (0–<5 years, 5–<10 years, 10–<15 years, 15–<20 years, and 21 years or more), with approximately 60 participants targeted in each category. Recruitment will also seek, as far as feasible, to reduce sex imbalance. Based on prior experience, men are expected to account for approximately 10% of available respondents, but recruitment will be adjusted as far as feasible toward a more balanced distribution.

Basic information collected in the survey will include age, sex, years of nursing experience, current principal clinical department, and main work setting.

Recruitment and survey administration will be conducted through [survey company name redacted] or a similar survey company. Eligibility will be restricted to persons who have continuously worked as nurses at a hospital or clinic in Japan during the one-year period immediately preceding the survey.

### 7. Protection of participants’ rights and personal information

The study data will not be used to identify individual participants and will be used only for research purposes. As described below, written signatures will not be collected, and personally identifying information such as names and home addresses will not be obtained. The personal characteristics collected will be limited to information used to describe and analyze the sample in publications, including sex, age, years of nursing experience, clinical department, and work setting.

Data obtained through the survey company will be anonymized by the platform (e.g., [survey company name redacted]), and the investigators will receive only anonymized data. The survey company will verify that participants are nurses.

Data will be stored under the responsibility of the principal investigator in a locked location for 10 years after the end of the study and then destroyed. Because some journals require or encourage publication of anonymized research data, participants will be informed before participation that anonymized response data may be made publicly available and consent will be obtained on that basis.

Participation will be based on voluntary consent. Participants will retain the right to refuse or discontinue participation at any time without disadvantage. Because only anonymized response data will be collected, participants will also be informed in advance that requests for deletion of their data after completion of the survey may not be possible.

### 8. Informed consent

Before the survey begins, participants will be shown the information contained in the “Request for Research Cooperation” document. Data collection will begin only for individuals who agree to participate under the stated conditions. Participation will be entirely voluntary, and declining participation will not result in any disadvantage.

Compensation will be processed within the survey company’s system; therefore, the investigators will not collect names, addresses, bank-account information, or other personal information required for payment. Accordingly, individuals cannot be identified from the response data. Consent will be obtained through a web-based consent form rather than a paper signature.

### 9. Measures to ensure participant safety

This study is a low-risk, non-invasive web-based psychological survey. The potential risks and safety information described in the “Request for Research Cooperation” will be presented before participation, and only individuals who consent will participate. Participants may discontinue the survey at any time. Participation is based solely on voluntary consent, and participants may refuse or discontinue participation at any time without disadvantage.

### 10. Additional information

#### 10.1 Provision of funding or equipment by companies or other organizations

Yes. A portion of the expenses required to conduct the study will be covered by commissioned research funds allocated to the Artificial Intelligence Research Center (AIRC), National Institute of Advanced Industrial Science and Technology (AIST), as described under “Breakdown of research expenses” below.

#### 10.2 Participant compensation

Yes. Participants who complete and submit all required data will receive compensation through the [survey company name redacted]. or equivalent web platform in accordance with the survey company’s rules. The estimated payment to the survey company was approximately JPY 1.2 million, subject to a quotation based on the number of participants and questions.

#### 10.3 Breakdown of research expenses

- Operating funds allocated to the principal investigator.
- JST PRESTO, Principal Investigator: R. Kagawa, Grant No. JPMJPR23I3, project title: “A diagnostic-support AI system for reducing cognitive digital disparities.”

#### 10.4 Conflicts of interest

None declared in the original ethics submission.

**Note:** This document is an English translation of the original Japanese study protocol submitted for ethical review. Some details of the survey as implemented differed from those described in the original protocol; the procedures actually used are described in the Methods section of the manuscript.

## Appendix B Details of Methods: Details of checking formats and scenario construction

The details of the checking formats specified in this study are presented below. Checking formats were organized according to three features: the number of people involved in checking, the number of checks within the relevant stage, and independence between checkers. The number of checks included both checking by another person and self-rechecking. Independence indicated whether multiple checkers each completed their check without referring to the other checker’s results.

Independence was specified only for the calculation stage. In the calculation stage, it was possible to define a format in which two people independently performed calculations from the same order information without referring to each other’s calculation results and then compared the results. In the preparation and administration stages, however, because the procedures involve physical tasks and actual administration to the patient, no corresponding independent checking format was specified within the scope of the scenarios used in this study.

### B.1 Calculation stage

The calculation stage involves calculating the dose, administration rate, concentration, dilution volume, and other values based on the physician’s orders or prescription information and, when necessary, converting these values into device settings (e.g., an infusion pump). Based on Dickinson et al. (2010), who categorized calculation-stage checking formats as independent checking, checking together, watching, and do and show, we added the “two-person sequential bidirectional” format described as a double-check method in Japanese patient-safety initiatives [9], as well as self-recalculation and a format with no recheck, yielding seven checking formats.

**Table B-1.**
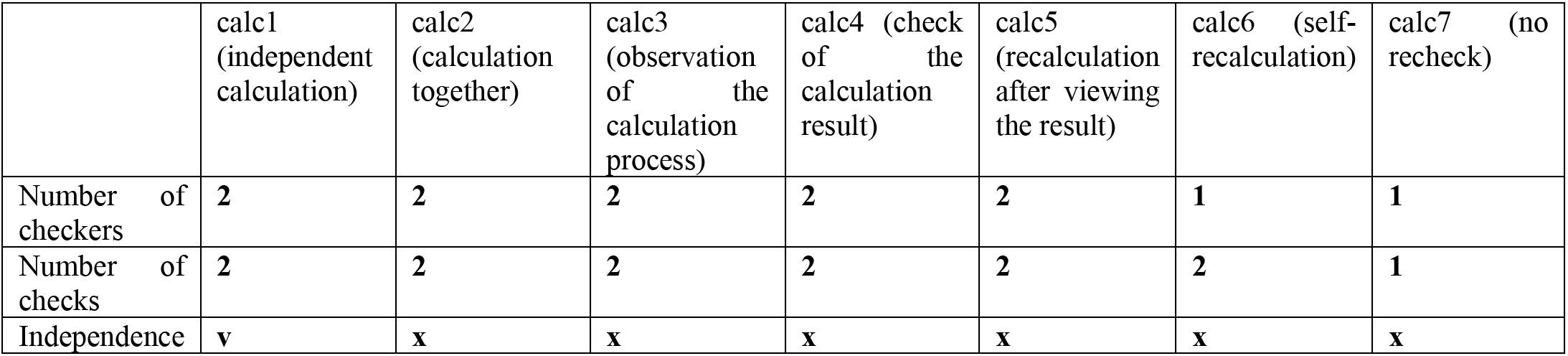
Checking formats in the calculation stage.

|  | calc1<br>(independent<br>calculation) | calc2<br>(calculation<br>together) | calc3<br>(observation<br>of the<br>calculation<br>process) | calc4 (check<br>of the<br>calculation<br>result) | calc5<br>(recalculation<br>after viewing<br>the result) | calc6 (self-recalculation) | calc7 (no<br>recheck) |
| --- | --- | --- | --- | --- | --- | --- | --- |
| Number of<br>checkers | 2 | 2 | 2 | 2 | 2 | 1 | 1 |
| Number of<br>checks | 2 | 2 | 2 | 2 | 2 | 2 | 1 |
| Independence | v | x | x | x | x | x | x |

The checking formats were as follows.

calc1 (independent calculation): Two people calculate independently without referring to each other’s calculation results and compare their results at the end (IDC).

calc2 (calculation together): Two people are present together and calculate together while verbally checking with each other or using similar methods.

calc3 (observation of the calculation process): One person calculates while the other observes the calculation process from beside them and identifies errors.

calc4 (check of the calculation result): One person presents the calculation result and the other checks that result.

calc5 (recalculation after viewing the result): After one person calculates, the other calculates while referring to that result and compares it with the original information, such as the physician’s order.

calc6 (self-recalculation): The same person calculates again and checks the calculation result.

calc7 (no recheck): One person calculates and no recheck is performed.

### B.2 Preparation stage

The preparation stage involves physical operations such as gathering medications, withdrawing medication from ampoules or vials, dilution, labeling, and setting up devices. Based on Dickinson et al. (2010), who categorized preparation-stage checking formats as done together and pre-prepared, we added self-rechecking and a format with no recheck, yielding four checking formats. Because the targets of checking in the scenarios included not only numerical information but also physical objects (the medication itself, labels, and equipment), we explicitly specified what was to be compared.

The checking formats were as follows.

prep1 (preparation and checking by two nurses together): Two people are present together and mutually check the medication name, dose, label, and related information while preparing the medication (or after preparation).

prep2 (second nurse checks after preparation): After one person prepares and checks the medication, the other compares the original information with the prepared medication itself (e.g., the syringe).

prep3 (self-recheck): One person prepares the medication and the same person checks it again. prep4 (no recheck): One person prepares the medication and no recheck is performed.

**Table B-2.**
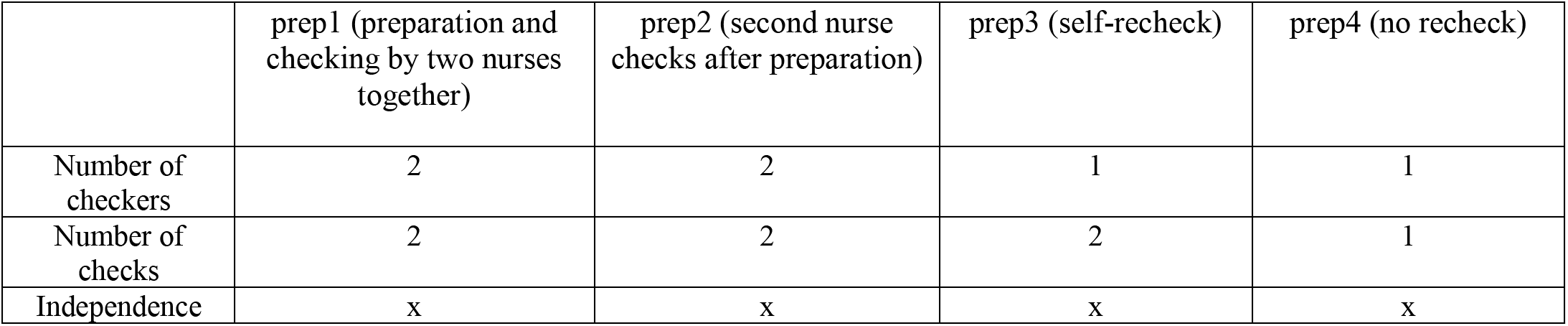
Checking formats in the preparation stage.

### B.3 Administration stage

The administration stage concerns the final act of administering medication to the patient and includes patient identification, checking the route and rate of administration, and initiating medication delivery. Based on Dickinson et al. (2010), who categorized administration-stage checking formats as one nurse at the bedside and two nurses at the bedside, we specified three checking formats, including self-rechecking by one person and a format with no recheck.

**Table B-3.** Checking formats in the administration stage.

|  | admin1 (pre-administration check by two nurses) | admin2 (self-recheck) | admin3 (no recheck) |
| --- | --- | --- | --- |
| Number of checkers | 2 | 1 | 1 |
| Number of checks | 2 | 2 | 1 |
| Independence | x | x | x |

The checking formats were as follows.

admin1 (pre-administration check by two nurses): Two people check the patient and administration conditions at the bedside, after which one person administers the medication.

admin2 (self-recheck): One person checks the patient and administration conditions at the bedside, rechecks the administration conditions before administration, and then administers the medication.

admin3 (no recheck): One person checks the patient and administration conditions and then administers the medication without a recheck.

### B.4 Exact wording used in the scenarios

Each scenario was constructed by combining one checking format from each of the calculation, preparation, and administration stages. The seven calculation formats, four preparation formats, and three administration formats yielded 84 scenarios in total (7 × 4 × 3). The code, name, and exact wording presented to participants for each checking format are shown in Tables B-4 to B-6.

**Table B-4.** Calculation-stage checking formats and scenario wording.

| Checking format | Scenario wording presented to participants |
| --- | --- |
| calc1 (independent calculation) | First, Nurse 1 calculates the medication dose alone based on the physician's orders and records the result. Nurse 2 then performs the same calculation using the same orders in a location away from Nurse 1 and records the result. The two nurses then compare their results and confirm that the numerical values match. |
| calc2 (calculation together) | First, Nurses 1 and 2 calculate the medication dose together based on the physician's orders, verbally checking with each other as they work, and record the result. The two nurses then compare their results and confirm that the numerical values match. |
| calc3 (observation of the calculation process) | First, Nurse 1 calculates the medication dose alone based on the physician's orders and records the result. While Nurse 1 is calculating, Nurse 2 observes the calculation process from beside Nurse 1 and checks for calculation errors. |
| calc4 (check of the calculation result) | First, Nurse 1 calculates the medication dose alone based on the physician's orders and records the result. Nurse 1 then tells Nurse 2 the calculation result, and Nurse 2 checks whether the calculation contains any errors. |
| calc5 (recalculation after viewing the result) | First, Nurse 1 calculates the medication dose alone based on the physician's orders and records the result. Nurse 2 then performs the calculation using Nurse 1's result as a reference and checks it against the physician's order to determine whether there are any calculation errors. |
| calc6 (self-recalculation) | First, Nurse 1 calculates the medication dose alone based on the physician's orders and records the result. Nurse 1 then performs the calculation again and checks whether there are any calculation errors. |
| calc7 (no recheck) | First, Nurse 1 calculates the medication dose alone based on the physician's orders and records the result. |

**Table B-5.** Preparation-stage checking formats and scenario wording.

| Checking format | Scenario wording presented to participants |
| --- | --- |
| prep1 (preparation and checking by two nurses together) | Next, Nurse 1 withdraws the medication from the vial into a syringe according to the physician's order. Nurses 1 and 2 then check the medication name and dose together, after which Nurse 1 applies the label. |
| prep2 (second nurse checks after preparation) | Next, Nurse 1 withdraws the medication from the vial into a syringe according to the physician's order and checks the medication name and dose. Nurse 1 then shows the syringe to Nurse 2 and states the medication name and dose. After Nurse 2 checks the contents of the syringe and the medication name and dose, Nurse 1 applies the label. |
| prep3 (self-recheck) | Next, Nurse 1 withdraws the medication from the vial into a syringe according to the physician's order and checks the medication name and dose. Nurse 1 then checks the medication name and dose again and applies the label. |
| prep4 (no recheck) | Next, Nurse 1 withdraws the medication from the vial into a syringe according to the physician's order and checks the medication name and dose, then applies the label. |

**Table B-6.** Administration-stage checking formats and scenario wording.

| Checking format | Scenario wording presented to participants |
| --- | --- |
| admin1 (pre-administration check by two nurses) | Finally, Nurses 1 and 2 go to the patient's bedside and verify the patient's identity against the medication information. They then check the medication name and dose together once more, after which Nurse 1 administers the prepared medication through the intravenous line. |
| admin2 (self-recheck) | Finally, Nurse 1 goes to the patient's bedside and verifies the patient's identity against the medication information. Nurse 1 then checks the medication name and dose once more and administers the prepared medication through the intravenous line. |
| admin3 (no recheck) | Finally, Nurse 1 goes to the patient's bedside and verifies the patient's identity against the medication information. Nurse 1 then administers the prepared medication through the intravenous line. |

## Appendix C Medication-administration scenarios

Note: calc/prep/admin are identification codes used only in this article and were not shown to participants.

### Scenario 1: calc1/prep1/admin1

The patient is a man in his 60s. A high-risk medication, Karicept, is to be administered intravenously to treat a condition requiring consideration of renal function. Nurses 1 and 2 are responsible for the patient.

First, Nurse 1 calculates the medication dose alone based on the physician’s orders and records the result. Nurse 2 then performs the same calculation using the same orders in a location away from Nurse 1 and records the result. The two nurses then compare their results and confirm that the numerical values match.

Next, Nurse 1 withdraws the medication from the vial into a syringe according to the physician’s order. Nurses 1 and 2 then check the medication name and dose together, after which Nurse 1 applies the label.

Finally, Nurses 1 and 2 go to the patient’s bedside and verify the patient’s identity against the medication information. They then check the medication name and dose together once more, after which Nurse 1 administers the prepared medication through the intravenous line.

### Scenario 2: calc1/prep1/admin2

The patient is a man in his 60s. A high-risk medication, Karicept, is to be administered intravenously to treat a condition requiring consideration of renal function. Nurses 1 and 2 are responsible for the patient.

First, Nurse 1 calculates the medication dose alone based on the physician’s orders and records the result. Nurse 2 then performs the same calculation using the same orders in a location away from Nurse 1 and records the result. The two nurses then compare their results and confirm that the numerical values match.

Next, Nurse 1 withdraws the medication from the vial into a syringe according to the physician’s order. Nurses 1 and 2 then check the medication name and dose together, after which Nurse 1 applies the label.

Finally, Nurse 1 goes to the patient’s bedside and verifies the patient’s identity against the medication information. Nurse 1 then checks the medication name and dose once more and administers the prepared medication through the intravenous line.

### Scenario 3: calc1/prep1/admin3

The patient is a man in his 60s. A high-risk medication, Karicept, is to be administered intravenously to treat a condition requiring consideration of renal function. Nurses 1 and 2 are responsible for the patient.

First, Nurse 1 calculates the medication dose alone based on the physician’s orders and records the result. Nurse 2 then performs the same calculation using the same orders in a location away from Nurse 1 and records the result. The two nurses then compare their results and confirm that the numerical values match.

Next, Nurse 1 withdraws the medication from the vial into a syringe according to the physician’s order. Nurses 1 and 2 then check the medication name and dose together, after which Nurse 1 applies the label.

Finally, Nurse 1 goes to the patient’s bedside and verifies the patient’s identity against the medication information. Nurse 1 then administers the prepared medication through the intravenous line.

### Scenario 4: calc1/prep2/admin1

The patient is a man in his 60s. A high-risk medication, Karicept, is to be administered intravenously to treat a condition requiring consideration of renal function. Nurses 1 and 2 are responsible for the patient.

First, Nurse 1 calculates the medication dose alone based on the physician’s orders and records the result. Nurse 2 then performs the same calculation using the same orders in a location away from Nurse 1 and records the result. The two nurses then compare their results and confirm that the numerical values match.

Next, Nurse 1 withdraws the medication from the vial into a syringe according to the physician’s order and checks the medication name and dose. Nurse 1 then shows the syringe to Nurse 2 and states the medication name and dose. After Nurse 2 checks the contents of the syringe and the medication name and dose, Nurse 1 applies the label.

Finally, Nurses 1 and 2 go to the patient’s bedside and verify the patient’s identity against the medication information. They then check the medication name and dose together once more, after which Nurse 1 administers the prepared medication through the intravenous line.

### Scenario 5: calc1/prep2/admin2

The patient is a man in his 60s. A high-risk medication, Karicept, is to be administered intravenously to treat a condition requiring consideration of renal function. Nurses 1 and 2 are responsible for the patient.

First, Nurse 1 calculates the medication dose alone based on the physician’s orders and records the result. Nurse 2 then performs the same calculation using the same orders in a location away from Nurse 1 and records the result. The two nurses then compare their results and confirm that the numerical values match.

Next, Nurse 1 withdraws the medication from the vial into a syringe according to the physician’s order and checks the medication name and dose. Nurse 1 then shows the syringe to Nurse 2 and states the medication name and dose. After Nurse 2 checks the contents of the syringe and the medication name and dose, Nurse 1 applies the label.

Finally, Nurse 1 goes to the patient’s bedside and verifies the patient’s identity against the medication information. Nurse 1 then checks the medication name and dose once more and administers the prepared medication through the intravenous line.

### Scenario 6: calc1/prep2/admin3

The patient is a man in his 60s. A high-risk medication, Karicept, is to be administered intravenously to treat a condition requiring consideration of renal function. Nurses 1 and 2 are responsible for the patient.

First, Nurse 1 calculates the medication dose alone based on the physician’s orders and records the result. Nurse 2 then performs the same calculation using the same orders in a location away from Nurse 1 and records the result. The two nurses then compare their results and confirm that the numerical values match.

Next, Nurse 1 withdraws the medication from the vial into a syringe according to the physician’s order and checks the medication name and dose. Nurse 1 then shows the syringe to Nurse 2 and states the medication name and dose. After Nurse 2 checks the contents of the syringe and the medication name and dose, Nurse 1 applies the label.

Finally, Nurse 1 goes to the patient’s bedside and verifies the patient’s identity against the medication information. Nurse 1 then administers the prepared medication through the intravenous line.

### Scenario 7: calc1/prep3/admin1

The patient is a man in his 60s. A high-risk medication, Karicept, is to be administered intravenously to treat a condition requiring consideration of renal function. Nurses 1 and 2 are responsible for the patient.

First, Nurse 1 calculates the medication dose alone based on the physician’s orders and records the result. Nurse 2 then performs the same calculation using the same orders in a location away from Nurse 1 and records the result. The two nurses then compare their results and confirm that the numerical values match.

Next, Nurse 1 withdraws the medication from the vial into a syringe according to the physician’s order and checks the medication name and dose. Nurse 1 then checks the medication name and dose again and applies the label.

Finally, Nurses 1 and 2 go to the patient’s bedside and verify the patient’s identity against the medication information. They then check the medication name and dose together once more, after which Nurse 1 administers the prepared medication through the intravenous line.

### Scenario 8: calc1/prep3/admin2

The patient is a man in his 60s. A high-risk medication, Karicept, is to be administered intravenously to treat a condition requiring consideration of renal function. Nurses 1 and 2 are responsible for the patient.

First, Nurse 1 calculates the medication dose alone based on the physician’s orders and records the result. Nurse 2 then performs the same calculation using the same orders in a location away from Nurse 1 and records the result. The two nurses then compare their results and confirm that the numerical values match.

Next, Nurse 1 withdraws the medication from the vial into a syringe according to the physician’s order and checks the medication name and dose. Nurse 1 then checks the medication name and dose again and applies the label.

Finally, Nurse 1 goes to the patient’s bedside and verifies the patient’s identity against the medication information. Nurse 1 then checks the medication name and dose once more and administers the prepared medication through the intravenous line.

### Scenario 9: calc1/prep3/admin3

The patient is a man in his 60s. A high-risk medication, Karicept, is to be administered intravenously to treat a condition requiring consideration of renal function. Nurses 1 and 2 are responsible for the patient.

First, Nurse 1 calculates the medication dose alone based on the physician’s orders and records the result. Nurse 2 then performs the same calculation using the same orders in a location away from Nurse 1 and records the result. The two nurses then compare their results and confirm that the numerical values match.

Next, Nurse 1 withdraws the medication from the vial into a syringe according to the physician’s order and checks the medication name and dose. Nurse 1 then checks the medication name and dose again and applies the label.

Finally, Nurse 1 goes to the patient’s bedside and verifies the patient’s identity against the medication information. Nurse 1 then administers the prepared medication through the intravenous line.

### Scenario 10: calc1/prep4/admin1

The patient is a man in his 60s. A high-risk medication, Karicept, is to be administered intravenously to treat a condition requiring consideration of renal function. Nurses 1 and 2 are responsible for the patient.

First, Nurse 1 calculates the medication dose alone based on the physician’s orders and records the result. Nurse 2 then performs the same calculation using the same orders in a location away from Nurse 1 and records the result. The two nurses then compare their results and confirm that the numerical values match.

Next, Nurse 1 withdraws the medication from the vial into a syringe according to the physician’s order and checks the medication name and dose, then applies the label.

Finally, Nurses 1 and 2 go to the patient’s bedside and verify the patient’s identity against the medication information. They then check the medication name and dose together once more, after which Nurse 1 administers the prepared medication through the intravenous line.

### Scenario 11: calc1/prep4/admin2

The patient is a man in his 60s. A high-risk medication, Karicept, is to be administered intravenously to treat a condition requiring consideration of renal function. Nurses 1 and 2 are responsible for the patient.

First, Nurse 1 calculates the medication dose alone based on the physician’s orders and records the result. Nurse 2 then performs the same calculation using the same orders in a location away from Nurse 1 and records the result. The two nurses then compare their results and confirm that the numerical values match.

Next, Nurse 1 withdraws the medication from the vial into a syringe according to the physician’s order and checks the medication name and dose, then applies the label.

Finally, Nurse 1 goes to the patient’s bedside and verifies the patient’s identity against the medication information. Nurse 1 then checks the medication name and dose once more and administers the prepared medication through the intravenous line.

### Scenario 12: calc1/prep4/admin3

The patient is a man in his 60s. A high-risk medication, Karicept, is to be administered intravenously to treat a condition requiring consideration of renal function. Nurses 1 and 2 are responsible for the patient.

First, Nurse 1 calculates the medication dose alone based on the physician’s orders and records the result. Nurse 2 then performs the same calculation using the same orders in a location away from Nurse 1 and records the result. The two nurses then compare their results and confirm that the numerical values match.

Next, Nurse 1 withdraws the medication from the vial into a syringe according to the physician’s order and checks the medication name and dose, then applies the label.

Finally, Nurse 1 goes to the patient’s bedside and verifies the patient’s identity against the medication information. Nurse 1 then administers the prepared medication through the intravenous line.

### Scenario 13: calc2/prep1/admin1

The patient is a man in his 60s. A high-risk medication, Karicept, is to be administered intravenously to treat a condition requiring consideration of renal function. Nurses 1 and 2 are responsible for the patient.

First, Nurses 1 and 2 calculate the medication dose together based on the physician’s orders, verbally checking with each other as they work, and record the result. The two nurses then compare their results and confirm that the numerical values match.

Next, Nurse 1 withdraws the medication from the vial into a syringe according to the physician’s order. Nurses 1 and 2 then check the medication name and dose together, after which Nurse 1 applies the label.

Finally, Nurses 1 and 2 go to the patient’s bedside and verify the patient’s identity against the medication information. They then check the medication name and dose together once more, after which Nurse 1 administers the prepared medication through the intravenous line.

### Scenario 14: calc2/prep1/admin2

The patient is a man in his 60s. A high-risk medication, Karicept, is to be administered intravenously to treat a condition requiring consideration of renal function. Nurses 1 and 2 are responsible for the patient.

First, Nurses 1 and 2 calculate the medication dose together based on the physician’s orders, verbally checking with each other as they work, and record the result. The two nurses then compare their results and confirm that the numerical values match.

Next, Nurse 1 withdraws the medication from the vial into a syringe according to the physician’s order. Nurses 1 and 2 then check the medication name and dose together, after which Nurse 1 applies the label.

Finally, Nurse 1 goes to the patient’s bedside and verifies the patient’s identity against the medication information. Nurse 1 then checks the medication name and dose once more and administers the prepared medication through the intravenous line.

### Scenario 15: calc2/prep1/admin3

The patient is a man in his 60s. A high-risk medication, Karicept, is to be administered intravenously to treat a condition requiring consideration of renal function. Nurses 1 and 2 are responsible for the patient.

First, Nurses 1 and 2 calculate the medication dose together based on the physician’s orders, verbally checking with each other as they work, and record the result. The two nurses then compare their results and confirm that the numerical values match.

Next, Nurse 1 withdraws the medication from the vial into a syringe according to the physician’s order. Nurses 1 and 2 then check the medication name and dose together, after which Nurse 1 applies the label.

Finally, Nurse 1 goes to the patient’s bedside and verifies the patient’s identity against the medication information. Nurse 1 then administers the prepared medication through the intravenous line.

### Scenario 16: calc2/prep2/admin1

The patient is a man in his 60s. A high-risk medication, Karicept, is to be administered intravenously to treat a condition requiring consideration of renal function. Nurses 1 and 2 are responsible for the patient.

First, Nurses 1 and 2 calculate the medication dose together based on the physician’s orders, verbally checking with each other as they work, and record the result. The two nurses then compare their results and confirm that the numerical values match.

Next, Nurse 1 withdraws the medication from the vial into a syringe according to the physician’s order and checks the medication name and dose. Nurse 1 then shows the syringe to Nurse 2 and states the medication name and dose. After Nurse 2 checks the contents of the syringe and the medication name and dose, Nurse 1 applies the label.

Finally, Nurses 1 and 2 go to the patient’s bedside and verify the patient’s identity against the medication information. They then check the medication name and dose together once more, after which Nurse 1 administers the prepared medication through the intravenous line.

### Scenario 17: calc2/prep2/admin2

The patient is a man in his 60s. A high-risk medication, Karicept, is to be administered intravenously to treat a condition requiring consideration of renal function. Nurses 1 and 2 are responsible for the patient.

First, Nurses 1 and 2 calculate the medication dose together based on the physician’s orders, verbally checking with each other as they work, and record the result. The two nurses then compare their results and confirm that the numerical values match.

Next, Nurse 1 withdraws the medication from the vial into a syringe according to the physician’s order and checks the medication name and dose. Nurse 1 then shows the syringe to Nurse 2 and states the medication name and dose. After Nurse 2 checks the contents of the syringe and the medication name and dose, Nurse 1 applies the label.

Finally, Nurse 1 goes to the patient’s bedside and verifies the patient’s identity against the medication information. Nurse 1 then checks the medication name and dose once more and administers the prepared medication through the intravenous line.

### Scenario 18: calc2/prep2/admin3

The patient is a man in his 60s. A high-risk medication, Karicept, is to be administered intravenously to treat a condition requiring consideration of renal function. Nurses 1 and 2 are responsible for the patient.

First, Nurses 1 and 2 calculate the medication dose together based on the physician’s orders, verbally checking with each other as they work, and record the result. The two nurses then compare their results and confirm that the numerical values match.

Next, Nurse 1 withdraws the medication from the vial into a syringe according to the physician’s order and checks the medication name and dose. Nurse 1 then shows the syringe to Nurse 2 and states the medication name and dose. After Nurse 2 checks the contents of the syringe and the medication name and dose, Nurse 1 applies the label.

Finally, Nurse 1 goes to the patient’s bedside and verifies the patient’s identity against the medication information. Nurse 1 then administers the prepared medication through the intravenous line.

### Scenario 19: calc2/prep3/admin1

The patient is a man in his 60s. A high-risk medication, Karicept, is to be administered intravenously to treat a condition requiring consideration of renal function. Nurses 1 and 2 are responsible for the patient.

First, Nurses 1 and 2 calculate the medication dose together based on the physician’s orders, verbally checking with each other as they work, and record the result. The two nurses then compare their results and confirm that the numerical values match.

Next, Nurse 1 withdraws the medication from the vial into a syringe according to the physician’s order and checks the medication name and dose. Nurse 1 then checks the medication name and dose again and applies the label.

Finally, Nurses 1 and 2 go to the patient’s bedside and verify the patient’s identity against the medication information. They then check the medication name and dose together once more, after which Nurse 1 administers the prepared medication through the intravenous line.

### Scenario 20: calc2/prep3/admin2

The patient is a man in his 60s. A high-risk medication, Karicept, is to be administered intravenously to treat a condition requiring consideration of renal function. Nurses 1 and 2 are responsible for the patient.

First, Nurses 1 and 2 calculate the medication dose together based on the physician’s orders, verbally checking with each other as they work, and record the result. The two nurses then compare their results and confirm that the numerical values match.

Next, Nurse 1 withdraws the medication from the vial into a syringe according to the physician’s order and checks the medication name and dose. Nurse 1 then checks the medication name and dose again and applies the label.

Finally, Nurse 1 goes to the patient’s bedside and verifies the patient’s identity against the medication information. Nurse 1 then checks the medication name and dose once more and administers the prepared medication through the intravenous line.

### Scenario 21: calc2/prep3/admin3

The patient is a man in his 60s. A high-risk medication, Karicept, is to be administered intravenously to treat a condition requiring consideration of renal function. Nurses 1 and 2 are responsible for the patient.

First, Nurses 1 and 2 calculate the medication dose together based on the physician’s orders, verbally checking with each other as they work, and record the result. The two nurses then compare their results and confirm that the numerical values match.

Next, Nurse 1 withdraws the medication from the vial into a syringe according to the physician’s order and checks the medication name and dose. Nurse 1 then checks the medication name and dose again and applies the label.

Finally, Nurse 1 goes to the patient’s bedside and verifies the patient’s identity against the medication information. Nurse 1 then administers the prepared medication through the intravenous line.

### Scenario 22: calc2/prep4/admin1

The patient is a man in his 60s. A high-risk medication, Karicept, is to be administered intravenously to treat a condition requiring consideration of renal function. Nurses 1 and 2 are responsible for the patient.

First, Nurses 1 and 2 calculate the medication dose together based on the physician’s orders, verbally checking with each other as they work, and record the result. The two nurses then compare their results and confirm that the numerical values match.

Next, Nurse 1 withdraws the medication from the vial into a syringe according to the physician’s order and checks the medication name and dose, then applies the label.

Finally, Nurses 1 and 2 go to the patient’s bedside and verify the patient’s identity against the medication information. They then check the medication name and dose together once more, after which Nurse 1 administers the prepared medication through the intravenous line.

### Scenario 23: calc2/prep4/admin2

The patient is a man in his 60s. A high-risk medication, Karicept, is to be administered intravenously to treat a condition requiring consideration of renal function. Nurses 1 and 2 are responsible for the patient.

First, Nurses 1 and 2 calculate the medication dose together based on the physician’s orders, verbally checking with each other as they work, and record the result. The two nurses then compare their results and confirm that the numerical values match.

Next, Nurse 1 withdraws the medication from the vial into a syringe according to the physician’s order and checks the medication name and dose, then applies the label.

Finally, Nurse 1 goes to the patient’s bedside and verifies the patient’s identity against the medication information. Nurse 1 then checks the medication name and dose once more and administers the prepared medication through the intravenous line.

### Scenario 24: calc2/prep4/admin3

The patient is a man in his 60s. A high-risk medication, Karicept, is to be administered intravenously to treat a condition requiring consideration of renal function. Nurses 1 and 2 are responsible for the patient.

First, Nurses 1 and 2 calculate the medication dose together based on the physician’s orders, verbally checking with each other as they work, and record the result. The two nurses then compare their results and confirm that the numerical values match.

Next, Nurse 1 withdraws the medication from the vial into a syringe according to the physician’s order and checks the medication name and dose, then applies the label.

Finally, Nurse 1 goes to the patient’s bedside and verifies the patient’s identity against the medication information. Nurse 1 then administers the prepared medication through the intravenous line.

### Scenario 25: calc3/prep1/admin1

The patient is a man in his 60s. A high-risk medication, Karicept, is to be administered intravenously to treat a condition requiring consideration of renal function. Nurses 1 and 2 are responsible for the patient.

First, Nurse 1 calculates the medication dose alone based on the physician’s orders and records the result. While Nurse 1 is calculating, Nurse 2 observes the calculation process from beside Nurse 1 and checks for calculation errors.

Next, Nurse 1 withdraws the medication from the vial into a syringe according to the physician’s order. Nurses 1 and 2 then check the medication name and dose together, after which Nurse 1 applies the label.

Finally, Nurses 1 and 2 go to the patient’s bedside and verify the patient’s identity against the medication information. They then check the medication name and dose together once more, after which Nurse 1 administers the prepared medication through the intravenous line.

### Scenario 26: calc3/prep1/admin2

The patient is a man in his 60s. A high-risk medication, Karicept, is to be administered intravenously to treat a condition requiring consideration of renal function. Nurses 1 and 2 are responsible for the patient.

First, Nurse 1 calculates the medication dose alone based on the physician’s orders and records the result. While Nurse 1 is calculating, Nurse 2 observes the calculation process from beside Nurse 1 and checks for calculation errors.

Next, Nurse 1 withdraws the medication from the vial into a syringe according to the physician’s order. Nurses 1 and 2 then check the medication name and dose together, after which Nurse 1 applies the label.

Finally, Nurse 1 goes to the patient’s bedside and verifies the patient’s identity against the medication information. Nurse 1 then checks the medication name and dose once more and administers the prepared medication through the intravenous line.

### Scenario 27: calc3/prep1/admin3

The patient is a man in his 60s. A high-risk medication, Karicept, is to be administered intravenously to treat a condition requiring consideration of renal function. Nurses 1 and 2 are responsible for the patient.

First, Nurse 1 calculates the medication dose alone based on the physician’s orders and records the result. While Nurse 1 is calculating, Nurse 2 observes the calculation process from beside Nurse 1 and checks for calculation errors.

Next, Nurse 1 withdraws the medication from the vial into a syringe according to the physician’s order. Nurses 1 and 2 then check the medication name and dose together, after which Nurse 1 applies the label.

Finally, Nurse 1 goes to the patient’s bedside and verifies the patient’s identity against the medication information. Nurse 1 then administers the prepared medication through the intravenous line.

### Scenario 28: calc3/prep2/admin1

The patient is a man in his 60s. A high-risk medication, Karicept, is to be administered intravenously to treat a condition requiring consideration of renal function. Nurses 1 and 2 are responsible for the patient.

First, Nurse 1 calculates the medication dose alone based on the physician’s orders and records the result. While Nurse 1 is calculating, Nurse 2 observes the calculation process from beside Nurse 1 and checks for calculation errors.

Next, Nurse 1 withdraws the medication from the vial into a syringe according to the physician’s order and checks the medication name and dose. Nurse 1 then shows the syringe to Nurse 2 and states the medication name and dose. After Nurse 2 checks the contents of the syringe and the medication name and dose, Nurse 1 applies the label.

Finally, Nurses 1 and 2 go to the patient’s bedside and verify the patient’s identity against the medication information. They then check the medication name and dose together once more, after which Nurse 1 administers the prepared medication through the intravenous line.

### Scenario 29: calc3/prep2/admin2

The patient is a man in his 60s. A high-risk medication, Karicept, is to be administered intravenously to treat a condition requiring consideration of renal function. Nurses 1 and 2 are responsible for the patient.

First, Nurse 1 calculates the medication dose alone based on the physician’s orders and records the result. While Nurse 1 is calculating, Nurse 2 observes the calculation process from beside Nurse 1 and checks for calculation errors.

Next, Nurse 1 withdraws the medication from the vial into a syringe according to the physician’s order and checks the medication name and dose. Nurse 1 then shows the syringe to Nurse 2 and states the medication name and dose. After Nurse 2 checks the contents of the syringe and the medication name and dose, Nurse 1 applies the label.

Finally, Nurse 1 goes to the patient’s bedside and verifies the patient’s identity against the medication information. Nurse 1 then checks the medication name and dose once more and administers the prepared medication through the intravenous line.

### Scenario 30: calc3/prep2/admin3

The patient is a man in his 60s. A high-risk medication, Karicept, is to be administered intravenously to treat a condition requiring consideration of renal function. Nurses 1 and 2 are responsible for the patient.

First, Nurse 1 calculates the medication dose alone based on the physician’s orders and records the result. While Nurse 1 is calculating, Nurse 2 observes the calculation process from beside Nurse 1 and checks for calculation errors.

Next, Nurse 1 withdraws the medication from the vial into a syringe according to the physician’s order and checks the medication name and dose. Nurse 1 then shows the syringe to Nurse 2 and states the medication name and dose. After Nurse 2 checks the contents of the syringe and the medication name and dose, Nurse 1 applies the label.

Finally, Nurse 1 goes to the patient’s bedside and verifies the patient’s identity against the medication information. Nurse 1 then administers the prepared medication through the intravenous line.

### Scenario 31: calc3/prep3/admin1

The patient is a man in his 60s. A high-risk medication, Karicept, is to be administered intravenously to treat a condition requiring consideration of renal function. Nurses 1 and 2 are responsible for the patient.

First, Nurse 1 calculates the medication dose alone based on the physician’s orders and records the result. While Nurse 1 is calculating, Nurse 2 observes the calculation process from beside Nurse 1 and checks for calculation errors.

Next, Nurse 1 withdraws the medication from the vial into a syringe according to the physician’s order and checks the medication name and dose. Nurse 1 then checks the medication name and dose again and applies the label.

Finally, Nurses 1 and 2 go to the patient’s bedside and verify the patient’s identity against the medication information. They then check the medication name and dose together once more, after which Nurse 1 administers the prepared medication through the intravenous line.

### Scenario 32: calc3/prep3/admin2

The patient is a man in his 60s. A high-risk medication, Karicept, is to be administered intravenously to treat a condition requiring consideration of renal function. Nurses 1 and 2 are responsible for the patient.

First, Nurse 1 calculates the medication dose alone based on the physician’s orders and records the result. While Nurse 1 is calculating, Nurse 2 observes the calculation process from beside Nurse 1 and checks for calculation errors.

Next, Nurse 1 withdraws the medication from the vial into a syringe according to the physician’s order and checks the medication name and dose. Nurse 1 then checks the medication name and dose again and applies the label.

Finally, Nurse 1 goes to the patient’s bedside and verifies the patient’s identity against the medication information. Nurse 1 then checks the medication name and dose once more and administers the prepared medication through the intravenous line.

### Scenario 33: calc3/prep3/admin3

The patient is a man in his 60s. A high-risk medication, Karicept, is to be administered intravenously to treat a condition requiring consideration of renal function. Nurses 1 and 2 are responsible for the patient.

First, Nurse 1 calculates the medication dose alone based on the physician’s orders and records the result. While Nurse 1 is calculating, Nurse 2 observes the calculation process from beside Nurse 1 and checks for calculation errors.

Next, Nurse 1 withdraws the medication from the vial into a syringe according to the physician’s order and checks the medication name and dose. Nurse 1 then checks the medication name and dose again and applies the label.

Finally, Nurse 1 goes to the patient’s bedside and verifies the patient’s identity against the medication information. Nurse 1 then administers the prepared medication through the intravenous line.

### Scenario 34: calc3/prep4/admin1

The patient is a man in his 60s. A high-risk medication, Karicept, is to be administered intravenously to treat a condition requiring consideration of renal function. Nurses 1 and 2 are responsible for the patient.

First, Nurse 1 calculates the medication dose alone based on the physician’s orders and records the result. While Nurse 1 is calculating, Nurse 2 observes the calculation process from beside Nurse 1 and checks for calculation errors.

Next, Nurse 1 withdraws the medication from the vial into a syringe according to the physician’s order and checks the medication name and dose, then applies the label.

Finally, Nurses 1 and 2 go to the patient’s bedside and verify the patient’s identity against the medication information. They then check the medication name and dose together once more, after which Nurse 1 administers the prepared medication through the intravenous line.

### Scenario 35: calc3/prep4/admin2

The patient is a man in his 60s. A high-risk medication, Karicept, is to be administered intravenously to treat a condition requiring consideration of renal function. Nurses 1 and 2 are responsible for the patient.

First, Nurse 1 calculates the medication dose alone based on the physician’s orders and records the result. While Nurse 1 is calculating, Nurse 2 observes the calculation process from beside Nurse 1 and checks for calculation errors.

Next, Nurse 1 withdraws the medication from the vial into a syringe according to the physician’s order and checks the medication name and dose, then applies the label.

Finally, Nurse 1 goes to the patient’s bedside and verifies the patient’s identity against the medication information. Nurse 1 then checks the medication name and dose once more and administers the prepared medication through the intravenous line.

### Scenario 36: calc3/prep4/admin3

The patient is a man in his 60s. A high-risk medication, Karicept, is to be administered intravenously to treat a condition requiring consideration of renal function. Nurses 1 and 2 are responsible for the patient.

First, Nurse 1 calculates the medication dose alone based on the physician’s orders and records the result. While Nurse 1 is calculating, Nurse 2 observes the calculation process from beside Nurse 1 and checks for calculation errors.

Next, Nurse 1 withdraws the medication from the vial into a syringe according to the physician’s order and checks the medication name and dose, then applies the label.

Finally, Nurse 1 goes to the patient’s bedside and verifies the patient’s identity against the medication information. Nurse 1 then administers the prepared medication through the intravenous line.

### Scenario 37: calc4/prep1/admin1

The patient is a man in his 60s. A high-risk medication, Karicept, is to be administered intravenously to treat a condition requiring consideration of renal function. Nurses 1 and 2 are responsible for the patient.

First, Nurse 1 calculates the medication dose alone based on the physician’s orders and records the result. Nurse 1 then tells Nurse 2 the calculation result, and Nurse 2 checks whether the calculation contains any errors.

Next, Nurse 1 withdraws the medication from the vial into a syringe according to the physician’s order. Nurses 1 and 2 then check the medication name and dose together, after which Nurse 1 applies the label.

Finally, Nurses 1 and 2 go to the patient’s bedside and verify the patient’s identity against the medication information. They then check the medication name and dose together once more, after which Nurse 1 administers the prepared medication through the intravenous line.

### Scenario 38: calc4/prep1/admin2

The patient is a man in his 60s. A high-risk medication, Karicept, is to be administered intravenously to treat a condition requiring consideration of renal function. Nurses 1 and 2 are responsible for the patient.

First, Nurse 1 calculates the medication dose alone based on the physician’s orders and records the result. Nurse 1 then tells Nurse 2 the calculation result, and Nurse 2 checks whether the calculation contains any errors.

Next, Nurse 1 withdraws the medication from the vial into a syringe according to the physician’s order. Nurses 1 and 2 then check the medication name and dose together, after which Nurse 1 applies the label.

Finally, Nurse 1 goes to the patient’s bedside and verifies the patient’s identity against the medication information. Nurse 1 then checks the medication name and dose once more and administers the prepared medication through the intravenous line.

### Scenario 39: calc4/prep1/admin3

The patient is a man in his 60s. A high-risk medication, Karicept, is to be administered intravenously to treat a condition requiring consideration of renal function. Nurses 1 and 2 are responsible for the patient.

First, Nurse 1 calculates the medication dose alone based on the physician’s orders and records the result. Nurse 1 then tells Nurse 2 the calculation result, and Nurse 2 checks whether the calculation contains any errors.

Next, Nurse 1 withdraws the medication from the vial into a syringe according to the physician’s order. Nurses 1 and 2 then check the medication name and dose together, after which Nurse 1 applies the label.

Finally, Nurse 1 goes to the patient’s bedside and verifies the patient’s identity against the medication information. Nurse 1 then administers the prepared medication through the intravenous line.

### Scenario 40: calc4/prep2/admin1

The patient is a man in his 60s. A high-risk medication, Karicept, is to be administered intravenously to treat a condition requiring consideration of renal function. Nurses 1 and 2 are responsible for the patient.

First, Nurse 1 calculates the medication dose alone based on the physician’s orders and records the result. Nurse 1 then tells Nurse 2 the calculation result, and Nurse 2 checks whether the calculation contains any errors.

Next, Nurse 1 withdraws the medication from the vial into a syringe according to the physician’s order and checks the medication name and dose. Nurse 1 then shows the syringe to Nurse 2 and states the medication name and dose. After Nurse 2 checks the contents of the syringe and the medication name and dose, Nurse 1 applies the label.

Finally, Nurses 1 and 2 go to the patient’s bedside and verify the patient’s identity against the medication information. They then check the medication name and dose together once more, after which Nurse 1 administers the prepared medication through the intravenous line.

### Scenario 41: calc4/prep2/admin2

The patient is a man in his 60s. A high-risk medication, Karicept, is to be administered intravenously to treat a condition requiring consideration of renal function. Nurses 1 and 2 are responsible for the patient.

First, Nurse 1 calculates the medication dose alone based on the physician’s orders and records the result. Nurse 1 then tells Nurse 2 the calculation result, and Nurse 2 checks whether the calculation contains any errors.

Next, Nurse 1 withdraws the medication from the vial into a syringe according to the physician’s order and checks the medication name and dose. Nurse 1 then shows the syringe to Nurse 2 and states the medication name and dose. After Nurse 2 checks the contents of the syringe and the medication name and dose, Nurse 1 applies the label.

Finally, Nurse 1 goes to the patient’s bedside and verifies the patient’s identity against the medication information. Nurse 1 then checks the medication name and dose once more and administers the prepared medication through the intravenous line.

### Scenario 42: calc4/prep2/admin3

The patient is a man in his 60s. A high-risk medication, Karicept, is to be administered intravenously to treat a condition requiring consideration of renal function. Nurses 1 and 2 are responsible for the patient.

First, Nurse 1 calculates the medication dose alone based on the physician’s orders and records the result. Nurse 1 then tells Nurse 2 the calculation result, and Nurse 2 checks whether the calculation contains any errors.

Next, Nurse 1 withdraws the medication from the vial into a syringe according to the physician’s order and checks the medication name and dose. Nurse 1 then shows the syringe to Nurse 2 and states the medication name and dose. After Nurse 2 checks the contents of the syringe and the medication name and dose, Nurse 1 applies the label.

Finally, Nurse 1 goes to the patient’s bedside and verifies the patient’s identity against the medication information. Nurse 1 then administers the prepared medication through the intravenous line.

### Scenario 43: calc4/prep3/admin1

The patient is a man in his 60s. A high-risk medication, Karicept, is to be administered intravenously to treat a condition requiring consideration of renal function. Nurses 1 and 2 are responsible for the patient.

First, Nurse 1 calculates the medication dose alone based on the physician’s orders and records the result. Nurse 1 then tells Nurse 2 the calculation result, and Nurse 2 checks whether the calculation contains any errors.

Next, Nurse 1 withdraws the medication from the vial into a syringe according to the physician’s order and checks the medication name and dose. Nurse 1 then checks the medication name and dose again and applies the label.

Finally, Nurses 1 and 2 go to the patient’s bedside and verify the patient’s identity against the medication information. They then check the medication name and dose together once more, after which Nurse 1 administers the prepared medication through the intravenous line.

### Scenario 44: calc4/prep3/admin2

The patient is a man in his 60s. A high-risk medication, Karicept, is to be administered intravenously to treat a condition requiring consideration of renal function. Nurses 1 and 2 are responsible for the patient.

First, Nurse 1 calculates the medication dose alone based on the physician’s orders and records the result. Nurse 1 then tells Nurse 2 the calculation result, and Nurse 2 checks whether the calculation contains any errors.

Next, Nurse 1 withdraws the medication from the vial into a syringe according to the physician’s order and checks the medication name and dose. Nurse 1 then checks the medication name and dose again and applies the label.

Finally, Nurse 1 goes to the patient’s bedside and verifies the patient’s identity against the medication information. Nurse 1 then checks the medication name and dose once more and administers the prepared medication through the intravenous line.

### Scenario 45: calc4/prep3/admin3

The patient is a man in his 60s. A high-risk medication, Karicept, is to be administered intravenously to treat a condition requiring consideration of renal function. Nurses 1 and 2 are responsible for the patient.

First, Nurse 1 calculates the medication dose alone based on the physician’s orders and records the result. Nurse 1 then tells Nurse 2 the calculation result, and Nurse 2 checks whether the calculation contains any errors.

Next, Nurse 1 withdraws the medication from the vial into a syringe according to the physician’s order and checks the medication name and dose. Nurse 1 then checks the medication name and dose again and applies the label.

Finally, Nurse 1 goes to the patient’s bedside and verifies the patient’s identity against the medication information. Nurse 1 then administers the prepared medication through the intravenous line.

### Scenario 46: calc4/prep4/admin1

The patient is a man in his 60s. A high-risk medication, Karicept, is to be administered intravenously to treat a condition requiring consideration of renal function. Nurses 1 and 2 are responsible for the patient.

First, Nurse 1 calculates the medication dose alone based on the physician’s orders and records the result. Nurse 1 then tells Nurse 2 the calculation result, and Nurse 2 checks whether the calculation contains any errors.

Next, Nurse 1 withdraws the medication from the vial into a syringe according to the physician’s order and checks the medication name and dose, then applies the label.

Finally, Nurses 1 and 2 go to the patient’s bedside and verify the patient’s identity against the medication information. They then check the medication name and dose together once more, after which Nurse 1 administers the prepared medication through the intravenous line.

### Scenario 47: calc4/prep4/admin2

The patient is a man in his 60s. A high-risk medication, Karicept, is to be administered intravenously to treat a condition requiring consideration of renal function. Nurses 1 and 2 are responsible for the patient.

First, Nurse 1 calculates the medication dose alone based on the physician’s orders and records the result. Nurse 1 then tells Nurse 2 the calculation result, and Nurse 2 checks whether the calculation contains any errors.

Next, Nurse 1 withdraws the medication from the vial into a syringe according to the physician’s order and checks the medication name and dose, then applies the label.

Finally, Nurse 1 goes to the patient’s bedside and verifies the patient’s identity against the medication information. Nurse 1 then checks the medication name and dose once more and administers the prepared medication through the intravenous line.

### Scenario 48: calc4/prep4/admin3

The patient is a man in his 60s. A high-risk medication, Karicept, is to be administered intravenously to treat a condition requiring consideration of renal function. Nurses 1 and 2 are responsible for the patient.

First, Nurse 1 calculates the medication dose alone based on the physician’s orders and records the result. Nurse 1 then tells Nurse 2 the calculation result, and Nurse 2 checks whether the calculation contains any errors.

Next, Nurse 1 withdraws the medication from the vial into a syringe according to the physician’s order and checks the medication name and dose, then applies the label.

Finally, Nurse 1 goes to the patient’s bedside and verifies the patient’s identity against the medication information. Nurse 1 then administers the prepared medication through the intravenous line.

### Scenario 49: calc5/prep1/admin1

The patient is a man in his 60s. A high-risk medication, Karicept, is to be administered intravenously to treat a condition requiring consideration of renal function. Nurses 1 and 2 are responsible for the patient.

First, Nurse 1 calculates the medication dose alone based on the physician’s orders and records the result. Nurse 2 then performs the calculation using Nurse 1’s result as a reference and checks it against the physician’s order to determine whether there are any calculation errors.

Next, Nurse 1 withdraws the medication from the vial into a syringe according to the physician’s order. Nurses 1 and 2 then check the medication name and dose together, after which Nurse 1 applies the label.

Finally, Nurses 1 and 2 go to the patient’s bedside and verify the patient’s identity against the medication information. They then check the medication name and dose together once more, after which Nurse 1 administers the prepared medication through the intravenous line.

### Scenario 50: calc5/prep1/admin2

The patient is a man in his 60s. A high-risk medication, Karicept, is to be administered intravenously to treat a condition requiring consideration of renal function. Nurses 1 and 2 are responsible for the patient.

First, Nurse 1 calculates the medication dose alone based on the physician’s orders and records the result. Nurse 2 then performs the calculation using Nurse 1’s result as a reference and checks it against the physician’s order to determine whether there are any calculation errors.

Next, Nurse 1 withdraws the medication from the vial into a syringe according to the physician’s order. Nurses 1 and 2 then check the medication name and dose together, after which Nurse 1 applies the label.

Finally, Nurse 1 goes to the patient’s bedside and verifies the patient’s identity against the medication information. Nurse 1 then checks the medication name and dose once more and administers the prepared medication through the intravenous line.

### Scenario 51: calc5/prep1/admin3

The patient is a man in his 60s. A high-risk medication, Karicept, is to be administered intravenously to treat a condition requiring consideration of renal function. Nurses 1 and 2 are responsible for the patient.

First, Nurse 1 calculates the medication dose alone based on the physician’s orders and records the result. Nurse 2 then performs the calculation using Nurse 1’s result as a reference and checks it against the physician’s order to determine whether there are any calculation errors.

Next, Nurse 1 withdraws the medication from the vial into a syringe according to the physician’s order. Nurses 1 and 2 then check the medication name and dose together, after which Nurse 1 applies the label.

Finally, Nurse 1 goes to the patient’s bedside and verifies the patient’s identity against the medication information. Nurse 1 then administers the prepared medication through the intravenous line.

### Scenario 52: calc5/prep2/admin1

The patient is a man in his 60s. A high-risk medication, Karicept, is to be administered intravenously to treat a condition requiring consideration of renal function. Nurses 1 and 2 are responsible for the patient.

First, Nurse 1 calculates the medication dose alone based on the physician’s orders and records the result. Nurse 2 then performs the calculation using Nurse 1’s result as a reference and checks it against the physician’s order to determine whether there are any calculation errors.

Next, Nurse 1 withdraws the medication from the vial into a syringe according to the physician’s order and checks the medication name and dose. Nurse 1 then shows the syringe to Nurse 2 and states the medication name and dose. After Nurse 2 checks the contents of the syringe and the medication name and dose, Nurse 1 applies the label.

Finally, Nurses 1 and 2 go to the patient’s bedside and verify the patient’s identity against the medication information. They then check the medication name and dose together once more, after which Nurse 1 administers the prepared medication through the intravenous line.

### Scenario 53: calc5/prep2/admin2

The patient is a man in his 60s. A high-risk medication, Karicept, is to be administered intravenously to treat a condition requiring consideration of renal function. Nurses 1 and 2 are responsible for the patient.

First, Nurse 1 calculates the medication dose alone based on the physician’s orders and records the result. Nurse 2 then performs the calculation using Nurse 1’s result as a reference and checks it against the physician’s order to determine whether there are any calculation errors.

Next, Nurse 1 withdraws the medication from the vial into a syringe according to the physician’s order and checks the medication name and dose. Nurse 1 then shows the syringe to Nurse 2 and states the medication name and dose. After Nurse 2 checks the contents of the syringe and the medication name and dose, Nurse 1 applies the label.

Finally, Nurse 1 goes to the patient’s bedside and verifies the patient’s identity against the medication information. Nurse 1 then checks the medication name and dose once more and administers the prepared medication through the intravenous line.

### Scenario 54: calc5/prep2/admin3

The patient is a man in his 60s. A high-risk medication, Karicept, is to be administered intravenously to treat a condition requiring consideration of renal function. Nurses 1 and 2 are responsible for the patient.

First, Nurse 1 calculates the medication dose alone based on the physician’s orders and records the result. Nurse 2 then performs the calculation using Nurse 1’s result as a reference and checks it against the physician’s order to determine whether there are any calculation errors.

Next, Nurse 1 withdraws the medication from the vial into a syringe according to the physician’s order and checks the medication name and dose. Nurse 1 then shows the syringe to Nurse 2 and states the medication name and dose. After Nurse 2 checks the contents of the syringe and the medication name and dose, Nurse 1 applies the label.

Finally, Nurse 1 goes to the patient’s bedside and verifies the patient’s identity against the medication information. Nurse 1 then administers the prepared medication through the intravenous line.

### Scenario 55: calc5/prep3/admin1

The patient is a man in his 60s. A high-risk medication, Karicept, is to be administered intravenously to treat a condition requiring consideration of renal function. Nurses 1 and 2 are responsible for the patient.

First, Nurse 1 calculates the medication dose alone based on the physician’s orders and records the result. Nurse 2 then performs the calculation using Nurse 1’s result as a reference and checks it against the physician’s order to determine whether there are any calculation errors.

Next, Nurse 1 withdraws the medication from the vial into a syringe according to the physician’s order and checks the medication name and dose. Nurse 1 then checks the medication name and dose again and applies the label.

Finally, Nurses 1 and 2 go to the patient’s bedside and verify the patient’s identity against the medication information. They then check the medication name and dose together once more, after which Nurse 1 administers the prepared medication through the intravenous line.

### Scenario 56: calc5/prep3/admin2

The patient is a man in his 60s. A high-risk medication, Karicept, is to be administered intravenously to treat a condition requiring consideration of renal function. Nurses 1 and 2 are responsible for the patient.

First, Nurse 1 calculates the medication dose alone based on the physician’s orders and records the result. Nurse 2 then performs the calculation using Nurse 1’s result as a reference and checks it against the physician’s order to determine whether there are any calculation errors.

Next, Nurse 1 withdraws the medication from the vial into a syringe according to the physician’s order and checks the medication name and dose. Nurse 1 then checks the medication name and dose again and applies the label.

Finally, Nurse 1 goes to the patient’s bedside and verifies the patient’s identity against the medication information. Nurse 1 then checks the medication name and dose once more and administers the prepared medication through the intravenous line.

### Scenario 57: calc5/prep3/admin3

The patient is a man in his 60s. A high-risk medication, Karicept, is to be administered intravenously to treat a condition requiring consideration of renal function. Nurses 1 and 2 are responsible for the patient.

First, Nurse 1 calculates the medication dose alone based on the physician’s orders and records the result. Nurse 2 then performs the calculation using Nurse 1’s result as a reference and checks it against the physician’s order to determine whether there are any calculation errors.

Next, Nurse 1 withdraws the medication from the vial into a syringe according to the physician’s order and checks the medication name and dose. Nurse 1 then checks the medication name and dose again and applies the label.

Finally, Nurse 1 goes to the patient’s bedside and verifies the patient’s identity against the medication information. Nurse 1 then administers the prepared medication through the intravenous line.

### Scenario 58: calc5/prep4/admin1

The patient is a man in his 60s. A high-risk medication, Karicept, is to be administered intravenously to treat a condition requiring consideration of renal function. Nurses 1 and 2 are responsible for the patient.

First, Nurse 1 calculates the medication dose alone based on the physician’s orders and records the result. Nurse 2 then performs the calculation using Nurse 1’s result as a reference and checks it against the physician’s order to determine whether there are any calculation errors.

Next, Nurse 1 withdraws the medication from the vial into a syringe according to the physician’s order and checks the medication name and dose, then applies the label.

Finally, Nurses 1 and 2 go to the patient’s bedside and verify the patient’s identity against the medication information. They then check the medication name and dose together once more, after which Nurse 1 administers the prepared medication through the intravenous line.

### Scenario 59: calc5/prep4/admin2

The patient is a man in his 60s. A high-risk medication, Karicept, is to be administered intravenously to treat a condition requiring consideration of renal function. Nurses 1 and 2 are responsible for the patient.

First, Nurse 1 calculates the medication dose alone based on the physician’s orders and records the result. Nurse 2 then performs the calculation using Nurse 1’s result as a reference and checks it against the physician’s order to determine whether there are any calculation errors.

Next, Nurse 1 withdraws the medication from the vial into a syringe according to the physician’s order and checks the medication name and dose, then applies the label.

Finally, Nurse 1 goes to the patient’s bedside and verifies the patient’s identity against the medication information. Nurse 1 then checks the medication name and dose once more and administers the prepared medication through the intravenous line.

### Scenario 60: calc5/prep4/admin3

The patient is a man in his 60s. A high-risk medication, Karicept, is to be administered intravenously to treat a condition requiring consideration of renal function. Nurses 1 and 2 are responsible for the patient.

First, Nurse 1 calculates the medication dose alone based on the physician’s orders and records the result. Nurse 2 then performs the calculation using Nurse 1’s result as a reference and checks it against the physician’s order to determine whether there are any calculation errors.

Next, Nurse 1 withdraws the medication from the vial into a syringe according to the physician’s order and checks the medication name and dose, then applies the label.

Finally, Nurse 1 goes to the patient’s bedside and verifies the patient’s identity against the medication information. Nurse 1 then administers the prepared medication through the intravenous line.

### Scenario 61: calc6/prep1/admin1

The patient is a man in his 60s. A high-risk medication, Karicept, is to be administered intravenously to treat a condition requiring consideration of renal function. Nurses 1 and 2 are responsible for the patient.

First, Nurse 1 calculates the medication dose alone based on the physician’s orders and records the result. Nurse 1 then performs the calculation again and checks whether there are any calculation errors.

Next, Nurse 1 withdraws the medication from the vial into a syringe according to the physician’s order. Nurses 1 and 2 then check the medication name and dose together, after which Nurse 1 applies the label.

Finally, Nurses 1 and 2 go to the patient’s bedside and verify the patient’s identity against the medication information. They then check the medication name and dose together once more, after which Nurse 1 administers the prepared medication through the intravenous line.

### Scenario 62: calc6/prep1/admin2

The patient is a man in his 60s. A high-risk medication, Karicept, is to be administered intravenously to treat a condition requiring consideration of renal function. Nurses 1 and 2 are responsible for the patient.

First, Nurse 1 calculates the medication dose alone based on the physician’s orders and records the result. Nurse 1 then performs the calculation again and checks whether there are any calculation errors.

Next, Nurse 1 withdraws the medication from the vial into a syringe according to the physician’s order. Nurses 1 and 2 then check the medication name and dose together, after which Nurse 1 applies the label.

Finally, Nurse 1 goes to the patient’s bedside and verifies the patient’s identity against the medication information. Nurse 1 then checks the medication name and dose once more and administers the prepared medication through the intravenous line.

### Scenario 63: calc6/prep1/admin3

The patient is a man in his 60s. A high-risk medication, Karicept, is to be administered intravenously to treat a condition requiring consideration of renal function. Nurses 1 and 2 are responsible for the patient.

First, Nurse 1 calculates the medication dose alone based on the physician’s orders and records the result. Nurse 1 then performs the calculation again and checks whether there are any calculation errors.

Next, Nurse 1 withdraws the medication from the vial into a syringe according to the physician’s order. Nurses 1 and 2 then check the medication name and dose together, after which Nurse 1 applies the label.

Finally, Nurse 1 goes to the patient’s bedside and verifies the patient’s identity against the medication information. Nurse 1 then administers the prepared medication through the intravenous line.

### Scenario 64: calc6/prep2/admin1

The patient is a man in his 60s. A high-risk medication, Karicept, is to be administered intravenously to treat a condition requiring consideration of renal function. Nurses 1 and 2 are responsible for the patient.

First, Nurse 1 calculates the medication dose alone based on the physician’s orders and records the result. Nurse 1 then performs the calculation again and checks whether there are any calculation errors.

Next, Nurse 1 withdraws the medication from the vial into a syringe according to the physician’s order and checks the medication name and dose. Nurse 1 then shows the syringe to Nurse 2 and states the medication name and dose. After Nurse 2 checks the contents of the syringe and the medication name and dose, Nurse 1 applies the label.

Finally, Nurses 1 and 2 go to the patient’s bedside and verify the patient’s identity against the medication information. They then check the medication name and dose together once more, after which Nurse 1 administers the prepared medication through the intravenous line.

### Scenario 65: calc6/prep2/admin2

The patient is a man in his 60s. A high-risk medication, Karicept, is to be administered intravenously to treat a condition requiring consideration of renal function. Nurses 1 and 2 are responsible for the patient.

First, Nurse 1 calculates the medication dose alone based on the physician’s orders and records the result. Nurse 1 then performs the calculation again and checks whether there are any calculation errors.

Next, Nurse 1 withdraws the medication from the vial into a syringe according to the physician’s order and checks the medication name and dose. Nurse 1 then shows the syringe to Nurse 2 and states the medication name and dose. After Nurse 2 checks the contents of the syringe and the medication name and dose, Nurse 1 applies the label.

Finally, Nurse 1 goes to the patient’s bedside and verifies the patient’s identity against the medication information. Nurse 1 then checks the medication name and dose once more and administers the prepared medication through the intravenous line.

### Scenario 66: calc6/prep2/admin3

The patient is a man in his 60s. A high-risk medication, Karicept, is to be administered intravenously to treat a condition requiring consideration of renal function. Nurses 1 and 2 are responsible for the patient.

First, Nurse 1 calculates the medication dose alone based on the physician’s orders and records the result. Nurse 1 then performs the calculation again and checks whether there are any calculation errors.

Next, Nurse 1 withdraws the medication from the vial into a syringe according to the physician’s order and checks the medication name and dose. Nurse 1 then shows the syringe to Nurse 2 and states the medication name and dose. After Nurse 2 checks the contents of the syringe and the medication name and dose, Nurse 1 applies the label.

Finally, Nurse 1 goes to the patient’s bedside and verifies the patient’s identity against the medication information. Nurse 1 then administers the prepared medication through the intravenous line.

### Scenario 67: calc6/prep3/admin1

The patient is a man in his 60s. A high-risk medication, Karicept, is to be administered intravenously to treat a condition requiring consideration of renal function. Nurses 1 and 2 are responsible for the patient.

First, Nurse 1 calculates the medication dose alone based on the physician’s orders and records the result. Nurse 1 then performs the calculation again and checks whether there are any calculation errors.

Next, Nurse 1 withdraws the medication from the vial into a syringe according to the physician’s order and checks the medication name and dose. Nurse 1 then checks the medication name and dose again and applies the label.

Finally, Nurses 1 and 2 go to the patient’s bedside and verify the patient’s identity against the medication information. They then check the medication name and dose together once more, after which Nurse 1 administers the prepared medication through the intravenous line.

### Scenario 68: calc6/prep3/admin2

The patient is a man in his 60s. A high-risk medication, Karicept, is to be administered intravenously to treat a condition requiring consideration of renal function. Nurses 1 and 2 are responsible for the patient.

First, Nurse 1 calculates the medication dose alone based on the physician’s orders and records the result. Nurse 1 then performs the calculation again and checks whether there are any calculation errors.

Next, Nurse 1 withdraws the medication from the vial into a syringe according to the physician’s order and checks the medication name and dose. Nurse 1 then checks the medication name and dose again and applies the label.

Finally, Nurse 1 goes to the patient’s bedside and verifies the patient’s identity against the medication information. Nurse 1 then checks the medication name and dose once more and administers the prepared medication through the intravenous line.

### Scenario 69: calc6/prep3/admin3

The patient is a man in his 60s. A high-risk medication, Karicept, is to be administered intravenously to treat a condition requiring consideration of renal function. Nurses 1 and 2 are responsible for the patient.

First, Nurse 1 calculates the medication dose alone based on the physician’s orders and records the result. Nurse 1 then performs the calculation again and checks whether there are any calculation errors.

Next, Nurse 1 withdraws the medication from the vial into a syringe according to the physician’s order and checks the medication name and dose. Nurse 1 then checks the medication name and dose again and applies the label.

Finally, Nurse 1 goes to the patient’s bedside and verifies the patient’s identity against the medication information. Nurse 1 then administers the prepared medication through the intravenous line.

### Scenario 70: calc6/prep4/admin1

The patient is a man in his 60s. A high-risk medication, Karicept, is to be administered intravenously to treat a condition requiring consideration of renal function. Nurses 1 and 2 are responsible for the patient.

First, Nurse 1 calculates the medication dose alone based on the physician’s orders and records the result. Nurse 1 then performs the calculation again and checks whether there are any calculation errors.

Next, Nurse 1 withdraws the medication from the vial into a syringe according to the physician’s order and checks the medication name and dose, then applies the label.

Finally, Nurses 1 and 2 go to the patient’s bedside and verify the patient’s identity against the medication information. They then check the medication name and dose together once more, after which Nurse 1 administers the prepared medication through the intravenous line.

### Scenario 71: calc6/prep4/admin2

The patient is a man in his 60s. A high-risk medication, Karicept, is to be administered intravenously to treat a condition requiring consideration of renal function. Nurses 1 and 2 are responsible for the patient.

First, Nurse 1 calculates the medication dose alone based on the physician’s orders and records the result. Nurse 1 then performs the calculation again and checks whether there are any calculation errors.

Next, Nurse 1 withdraws the medication from the vial into a syringe according to the physician’s order and checks the medication name and dose, then applies the label.

Finally, Nurse 1 goes to the patient’s bedside and verifies the patient’s identity against the medication information. Nurse 1 then checks the medication name and dose once more and administers the prepared medication through the intravenous line.

### Scenario 72: calc6/prep4/admin3

The patient is a man in his 60s. A high-risk medication, Karicept, is to be administered intravenously to treat a condition requiring consideration of renal function. Nurses 1 and 2 are responsible for the patient.

First, Nurse 1 calculates the medication dose alone based on the physician’s orders and records the result. Nurse 1 then performs the calculation again and checks whether there are any calculation errors.

Next, Nurse 1 withdraws the medication from the vial into a syringe according to the physician’s order and checks the medication name and dose, then applies the label.

Finally, Nurse 1 goes to the patient’s bedside and verifies the patient’s identity against the medication information. Nurse 1 then administers the prepared medication through the intravenous line.

### Scenario 73: calc7/prep1/admin1

The patient is a man in his 60s. A high-risk medication, Karicept, is to be administered intravenously to treat a condition requiring consideration of renal function. Nurses 1 and 2 are responsible for the patient.

First, Nurse 1 calculates the medication dose alone based on the physician’s orders and records the result.

Next, Nurse 1 withdraws the medication from the vial into a syringe according to the physician’s order. Nurses 1 and 2 then check the medication name and dose together, after which Nurse 1 applies the label.

Finally, Nurses 1 and 2 go to the patient’s bedside and verify the patient’s identity against the medication information. They then check the medication name and dose together once more, after which Nurse 1 administers the prepared medication through the intravenous line.

### Scenario 74: calc7/prep1/admin2

The patient is a man in his 60s. A high-risk medication, Karicept, is to be administered intravenously to treat a condition requiring consideration of renal function. Nurses 1 and 2 are responsible for the patient.

First, Nurse 1 calculates the medication dose alone based on the physician’s orders and records the result.

Next, Nurse 1 withdraws the medication from the vial into a syringe according to the physician’s order. Nurses 1 and 2 then check the medication name and dose together, after which Nurse 1 applies the label.

Finally, Nurse 1 goes to the patient’s bedside and verifies the patient’s identity against the medication information. Nurse 1 then checks the medication name and dose once more and administers the prepared medication through the intravenous line.

### Scenario 75: calc7/prep1/admin3

The patient is a man in his 60s. A high-risk medication, Karicept, is to be administered intravenously to treat a condition requiring consideration of renal function. Nurses 1 and 2 are responsible for the patient.

First, Nurse 1 calculates the medication dose alone based on the physician’s orders and records the result.

Next, Nurse 1 withdraws the medication from the vial into a syringe according to the physician’s order. Nurses 1 and 2 then check the medication name and dose together, after which Nurse 1 applies the label.

Finally, Nurse 1 goes to the patient’s bedside and verifies the patient’s identity against the medication information. Nurse 1 then administers the prepared medication through the intravenous line.

### Scenario 76: calc7/prep2/admin1

The patient is a man in his 60s. A high-risk medication, Karicept, is to be administered intravenously to treat a condition requiring consideration of renal function. Nurses 1 and 2 are responsible for the patient.

First, Nurse 1 calculates the medication dose alone based on the physician’s orders and records the result.

Next, Nurse 1 withdraws the medication from the vial into a syringe according to the physician’s order and checks the medication name and dose. Nurse 1 then shows the syringe to Nurse 2 and states the medication name and dose. After Nurse 2 checks the contents of the syringe and the medication name and dose, Nurse 1 applies the label.

Finally, Nurses 1 and 2 go to the patient’s bedside and verify the patient’s identity against the medication information. They then check the medication name and dose together once more, after which Nurse 1 administers the prepared medication through the intravenous line.

### Scenario 77: calc7/prep2/admin2

The patient is a man in his 60s. A high-risk medication, Karicept, is to be administered intravenously to treat a condition requiring consideration of renal function. Nurses 1 and 2 are responsible for the patient.

First, Nurse 1 calculates the medication dose alone based on the physician’s orders and records the result.

Next, Nurse 1 withdraws the medication from the vial into a syringe according to the physician’s order and checks the medication name and dose. Nurse 1 then shows the syringe to Nurse 2 and states the medication name and dose. After Nurse 2 checks the contents of the syringe and the medication name and dose, Nurse 1 applies the label.

Finally, Nurse 1 goes to the patient’s bedside and verifies the patient’s identity against the medication information. Nurse 1 then checks the medication name and dose once more and administers the prepared medication through the intravenous line.

### Scenario 78: calc7/prep2/admin3

The patient is a man in his 60s. A high-risk medication, Karicept, is to be administered intravenously to treat a condition requiring consideration of renal function. Nurses 1 and 2 are responsible for the patient.

First, Nurse 1 calculates the medication dose alone based on the physician’s orders and records the result.

Next, Nurse 1 withdraws the medication from the vial into a syringe according to the physician’s order and checks the medication name and dose. Nurse 1 then shows the syringe to Nurse 2 and states the medication name and dose. After Nurse 2 checks the contents of the syringe and the medication name and dose, Nurse 1 applies the label.

Finally, Nurse 1 goes to the patient’s bedside and verifies the patient’s identity against the medication information. Nurse 1 then administers the prepared medication through the intravenous line.

### Scenario 79: calc7/prep3/admin1

The patient is a man in his 60s. A high-risk medication, Karicept, is to be administered intravenously to treat a condition requiring consideration of renal function. Nurses 1 and 2 are responsible for the patient.

First, Nurse 1 calculates the medication dose alone based on the physician’s orders and records the result.

Next, Nurse 1 withdraws the medication from the vial into a syringe according to the physician’s order and checks the medication name and dose. Nurse 1 then checks the medication name and dose again and applies the label.

Finally, Nurses 1 and 2 go to the patient’s bedside and verify the patient’s identity against the medication information. They then check the medication name and dose together once more, after which Nurse 1 administers the prepared medication through the intravenous line.

### Scenario 80: calc7/prep3/admin2

The patient is a man in his 60s. A high-risk medication, Karicept, is to be administered intravenously to treat a condition requiring consideration of renal function. Nurses 1 and 2 are responsible for the patient.

First, Nurse 1 calculates the medication dose alone based on the physician’s orders and records the result.

Next, Nurse 1 withdraws the medication from the vial into a syringe according to the physician’s order and checks the medication name and dose. Nurse 1 then checks the medication name and dose again and applies the label.

Finally, Nurse 1 goes to the patient’s bedside and verifies the patient’s identity against the medication information. Nurse 1 then checks the medication name and dose once more and administers the prepared medication through the intravenous line.

### Scenario 81: calc7/prep3/admin3

The patient is a man in his 60s. A high-risk medication, Karicept, is to be administered intravenously to treat a condition requiring consideration of renal function. Nurses 1 and 2 are responsible for the patient.

First, Nurse 1 calculates the medication dose alone based on the physician’s orders and records the result.

Next, Nurse 1 withdraws the medication from the vial into a syringe according to the physician’s order and checks the medication name and dose. Nurse 1 then checks the medication name and dose again and applies the label.

Finally, Nurse 1 goes to the patient’s bedside and verifies the patient’s identity against the medication information. Nurse 1 then administers the prepared medication through the intravenous line.

### Scenario 82: calc7/prep4/admin1

The patient is a man in his 60s. A high-risk medication, Karicept, is to be administered intravenously to treat a condition requiring consideration of renal function. Nurses 1 and 2 are responsible for the patient.

First, Nurse 1 calculates the medication dose alone based on the physician’s orders and records the result.

Next, Nurse 1 withdraws the medication from the vial into a syringe according to the physician’s order and checks the medication name and dose, then applies the label.

Finally, Nurses 1 and 2 go to the patient’s bedside and verify the patient’s identity against the medication information. They then check the medication name and dose together once more, after which Nurse 1 administers the prepared medication through the intravenous line.

### Scenario 83: calc7/prep4/admin2

The patient is a man in his 60s. A high-risk medication, Karicept, is to be administered intravenously to treat a condition requiring consideration of renal function. Nurses 1 and 2 are responsible for the patient.

First, Nurse 1 calculates the medication dose alone based on the physician’s orders and records the result.

Next, Nurse 1 withdraws the medication from the vial into a syringe according to the physician’s order and checks the medication name and dose, then applies the label.

Finally, Nurse 1 goes to the patient’s bedside and verifies the patient’s identity against the medication information. Nurse 1 then checks the medication name and dose once more and administers the prepared medication through the intravenous line.

### Scenario 84: calc7/prep4/admin3

The patient is a man in his 60s. A high-risk medication, Karicept, is to be administered intravenously to treat a condition requiring consideration of renal function. Nurses 1 and 2 are responsible for the patient.

First, Nurse 1 calculates the medication dose alone based on the physician’s orders and records the result.

Next, Nurse 1 withdraws the medication from the vial into a syringe according to the physician’s order and checks the medication name and dose, then applies the label.

Finally, Nurse 1 goes to the patient’s bedside and verifies the patient’s identity against the medication information. Nurse 1 then administers the prepared medication through the intravenous line.

## Appendix D Details of Results

### D.1 Analysis using linear mixed-effects models

#### D.1.1 Model construction

In this section, hierarchical models were used to examine how features of checking procedures (independence, number of checkers, and number of checks) and respondent characteristics were associated with desirability (Q1) and implementation frequency (Q2 and Q3), while simultaneously estimating the mutually adjusted effects of scenario-level checking-procedure features and respondent-level characteristics. Because the data had a repeated-measures structure in which the same respondent rated multiple scenarios, linear mixed-effects models (LMMs) including respondent ID as a random intercept were used.

Hereafter, respondent is denoted by *i*, observation (scenario) by *j*, and the 7-point rating by *y_i_*_j_. Predictors were defined separately as scenario-level procedure features, Z*_i_*_j_, and respondent-level characteristics, X*_i_*. Procedure features Z*_i_*_j_consisted of independence, number of checkers, and number of checks. Respondent characteristics X*_i_* consisted of number of beds, years of experience, frequency of non-high-risk medication administration, and frequency of high-risk medication administration.

**Table D-1.**
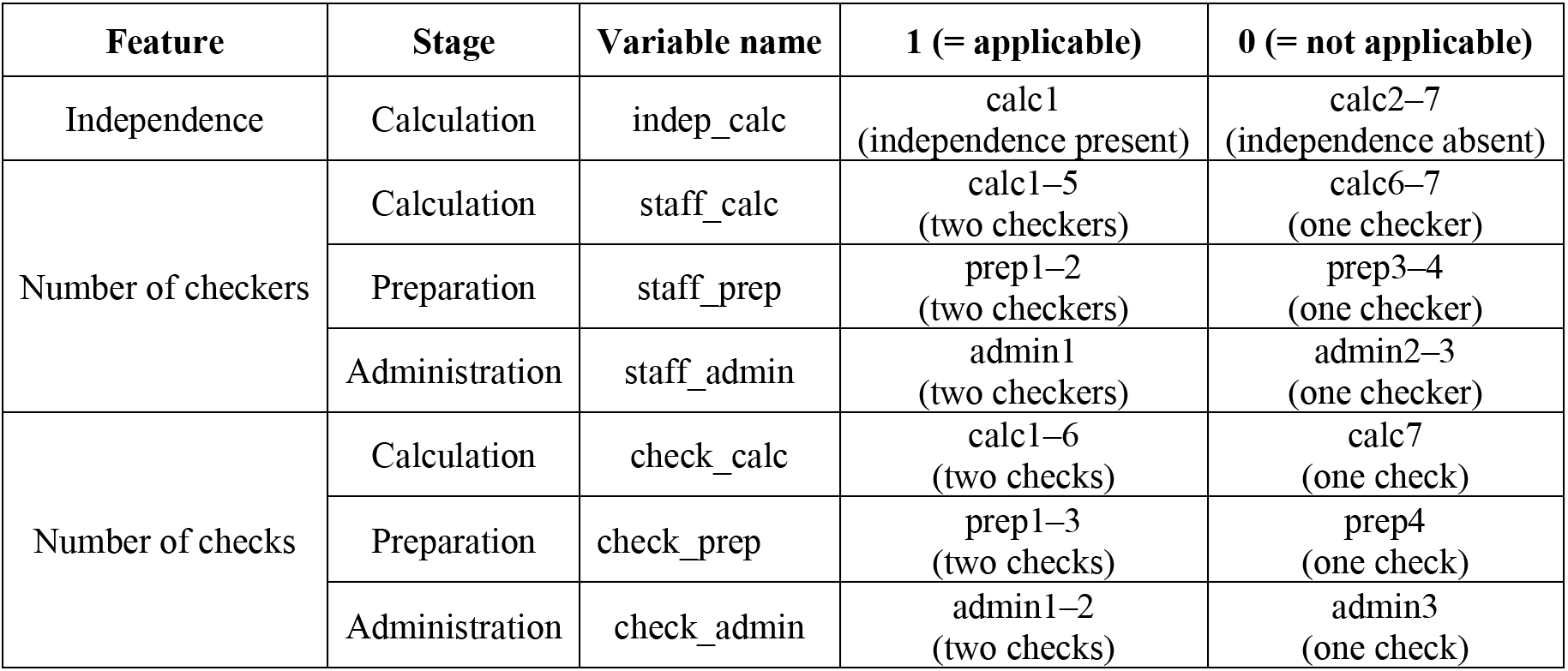
Definitions of procedure features and correspondence with scenario components.

**Table D-2.** Definitions of respondent-characteristic categories.

| Variable | Code | Category |
| --- | --- | --- |
| beds (number of beds) | 1 | No inpatient beds (0 beds) |
| beds (number of beds) | 2 | 1–19 beds |
| beds (number of beds) | 3 | 20–99 beds |
| beds (number of beds) | 4 | 100–199 beds |
| beds (number of beds) | 5 | 200–299 beds |
| beds (number of beds) | 6 | 300–499 beds |
| beds (number of beds) | 7 | ≥500 beds |
| beds (number of beds) | 8 | Other/unknown |
| tenure (years of nursing experience) | 1 | <5 years |
| tenure (years of nursing experience) | 2 | 5 to <10 years |
| tenure (years of nursing experience) | 3 | 10 to <15 years |
| tenure (years of nursing experience) | 4 | 15 to <20 years |
| tenure (years of nursing experience) | 5 | ≥20 years |
| freq_reg (frequency of non-high-risk medication administration) | 1 | Never |
| freq_reg (frequency of non-high-risk medication administration) | 2 | Rarely (<1/month) |
| freq_reg (frequency of non-high-risk medication administration) | 3 | Several times/month (<1/week) |
| freq_reg (frequency of non-high-risk medication administration) | 4 | Several times/week (about 1 to several/week) |
| freq_reg (frequency of non-high-risk medication administration) | 5 | Almost daily (on nearly all working days) |
| freq_high (frequency of high-risk medication administration) | 1 | Never |
| freq_high (frequency of high-risk medication administration) | 2 | Rarely (<1/month) |
| freq_high (frequency of high-risk medication administration) | 3 | Several times/month (<1/week) |
| freq_high (frequency of high-risk medication administration) | 4 | Several times/week (about 1 to several/week) |
| freq_high (frequency of high-risk medication administration) | 5 | Almost daily (on nearly all working days) |

To avoid an excessive number of categories, respondent characteristics were entered as ordinal variables (beds_ord, tenure_ord, freq_reg_ord, freq_high_ord). Because the category coded as beds=8 could not be meaningfully ordered by facility size, it was treated separately as the dummy variable beds_unk (applicable=1, otherwise=0).

For each of Q1–Q3, three models were estimated. Model 1 estimated the main effects of checking-procedure features. Model 2 added respondent characteristics to estimate main effects adjusted for these characteristics. Model 3 introduced interaction terms to examine whether the association between checking-procedure features and the outcomes varied by respondent characteristics. However, including interactions between every feature and every respondent characteristic would sharply increase the number of parameters and could destabilize estimation and reduce interpretability. Therefore, Model 3 focused on independence, which was a primary focus of this study and a defining feature of independent double-checks, and restricted interactions to independence × respondent-characteristics terms.

Model 1 (features) was defined as follows.

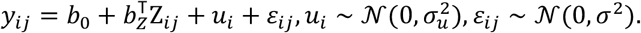

Model 2 (features + characteristics) was defined as follows.

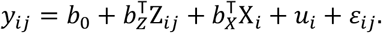

Model 3 (features + characteristics + interactions: independence × characteristics) included independence as well as its interactions with respondent characteristics and was specified as follows.

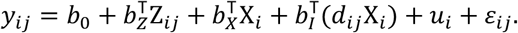

The fixed-effect coefficients *b* are unstandardized coefficients and represent the average change in the outcome variable, in points, associated with a one-unit change in the predictor while holding the other variables constant. *u_i_* represents the between-respondent difference in baseline level (random intercept).

Before model estimation, multicollinearity among independent variables was examined. First, variance inflation factors (VIFs) were calculated for Model 2, in which the main effects and respondent characteristics were entered simultaneously. Because each predictor in the present model had 1 degree of freedom, the calculated generalized variance inflation factor (GVIF^1/(2df)^) was squared to convert it to the usual VIF scale. The maximum VIF was 4.40, below the commonly used threshold of 5, indicating no serious multicollinearity. In the Q2 and Q3 models, no respondents selected “other/unknown” for number of beds in the clinical setting, making the corresponding dummy variable constant; this variable was therefore excluded from those analyses. Next, Model 3 added interaction terms between independence and respondent characteristics. Because models containing interaction terms may induce correlations between main effects and interaction terms, Model 3 was evaluated using the VIF values together with convergence status and changes in coefficients and standard errors. These checks indicated that no serious problems affected the estimates.

### D.2 Rating patterns by checking procedure

For Q1 (desirability of the checking procedure), Q2 (implementation frequency for high-risk medications), and Q3 (implementation frequency for non-high-risk medications), rating patterns were also examined for each checking procedure within the calculation, preparation, and administration stages. Differences in ratings among checking procedures within each stage were examined using Friedman tests, and the response distributions across rating categories were described.

#### D.2.1 Tests of differences in ratings among procedures

In this study, the same respondent evaluated multiple checking procedures within each stage. Therefore, for each respondent, the mean rating of scenarios containing each checking procedure was calculated, and differences in ratings among checking procedures for Q1–Q3 were examined separately within the calculation, preparation, and administration stages using Friedman tests. Kendall’s W was calculated as the effect size.

**Table D-3.** Friedman tests of differences in ratings among checking procedures.

| Question | Stage | N | $\chi^2$ (df) | p value | Effect size (W) |
| --- | --- | --- | --- | --- | --- |
| Q1 | Calculation (calc) | 265 | 863.00 (6) | <0.001 | 0.543 |
| Q1 | Preparation (prep) | 265 | 519.84 (3) | <0.001 | 0.654 |
| Q1 | Administration (admin) | 265 | 290.02 (2) | <0.001 | 0.547 |
| Q2 | Calculation (calc) | 222 | 384.02 (6) | <0.001 | 0.288 |
| Q2 | Preparation (prep) | 222 | 259.30 (3) | <0.001 | 0.389 |
| Q2 | Administration (admin) | 222 | 35.27 (2) | <0.001 | 0.079 |
| Q3 | Calculation (calc) | 255 | 132.52 (6) | <0.001 | 0.087 |
| Q3 | Preparation (prep) | 255 | 90.29 (3) | <0.001 | 0.118 |
| Q3 | Administration (admin) | 255 | 10.71 (2) | 0.005 | 0.021 |

The results are shown in Table D-3. For Q1, significant differences among checking procedures were observed in all stages: calculation (χ^2^ (6)=863.00, p<0.001, W=0.543), preparation ( χ^2^ (3)=519.84, p<0.001, W=0.654), and administration (χ^2^(2)=290.02, p<0.001, W=0.547). For Q2, significant differences were likewise observed for calculation (χ^2^(6)=384.02, p<0.001, W=0.288), preparation (χ^2^(3)=259.30, p<0.001, W=0.389), and administration (χ^2^(2)=35.27, p<0.001, W=0.079). For Q3, significant differences were also observed for calculation ( χ^2^ (6)=132.52, p<0.001, W=0.087), preparation (χ^2^(3)=90.29, p<0.001, W=0.118), and administration (χ^2^(2)=10.71, p=0.005, W=0.021). Effect sizes were largest for Q1 and were generally smaller for Q2 and Q3.

#### D.2.2 Visualization of item-level response patterns

The Friedman tests examined differences in ratings among checking procedures within each stage. However, they do not show the distribution of ratings for each individual checking procedure. Therefore, for Q1–Q3, we calculated the percentage distribution across rating categories for each checking procedure.

##### (1) Calculation stage

For Q1, the proportions of ratings 5–7 were relatively high for calc1, whereas the proportions of ratings 1–3 were high for calc6 and calc7. Calc2–calc5 were intermediate. For Q2, the distributions for calc1–calc5 were relatively similar, whereas calc6 and calc7 had higher proportions of low ratings. A similar pattern was observed for Q3, although differences in the distributions among checking procedures, including calc6 and calc7, were smaller than for Q1.

**Table D-4a.**
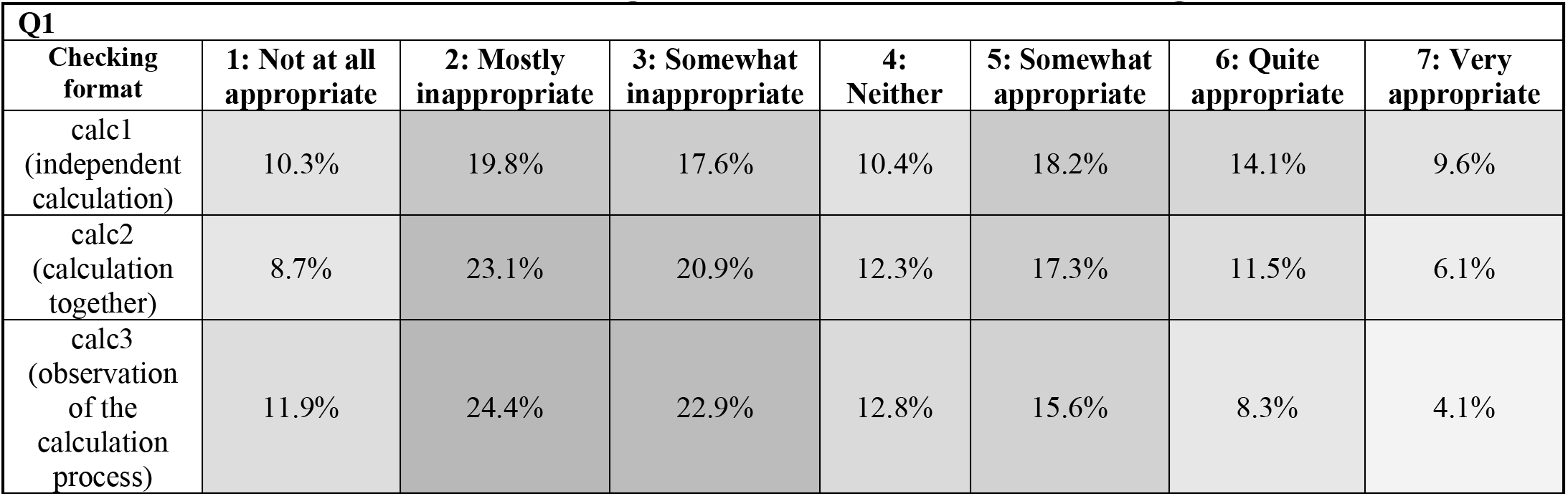
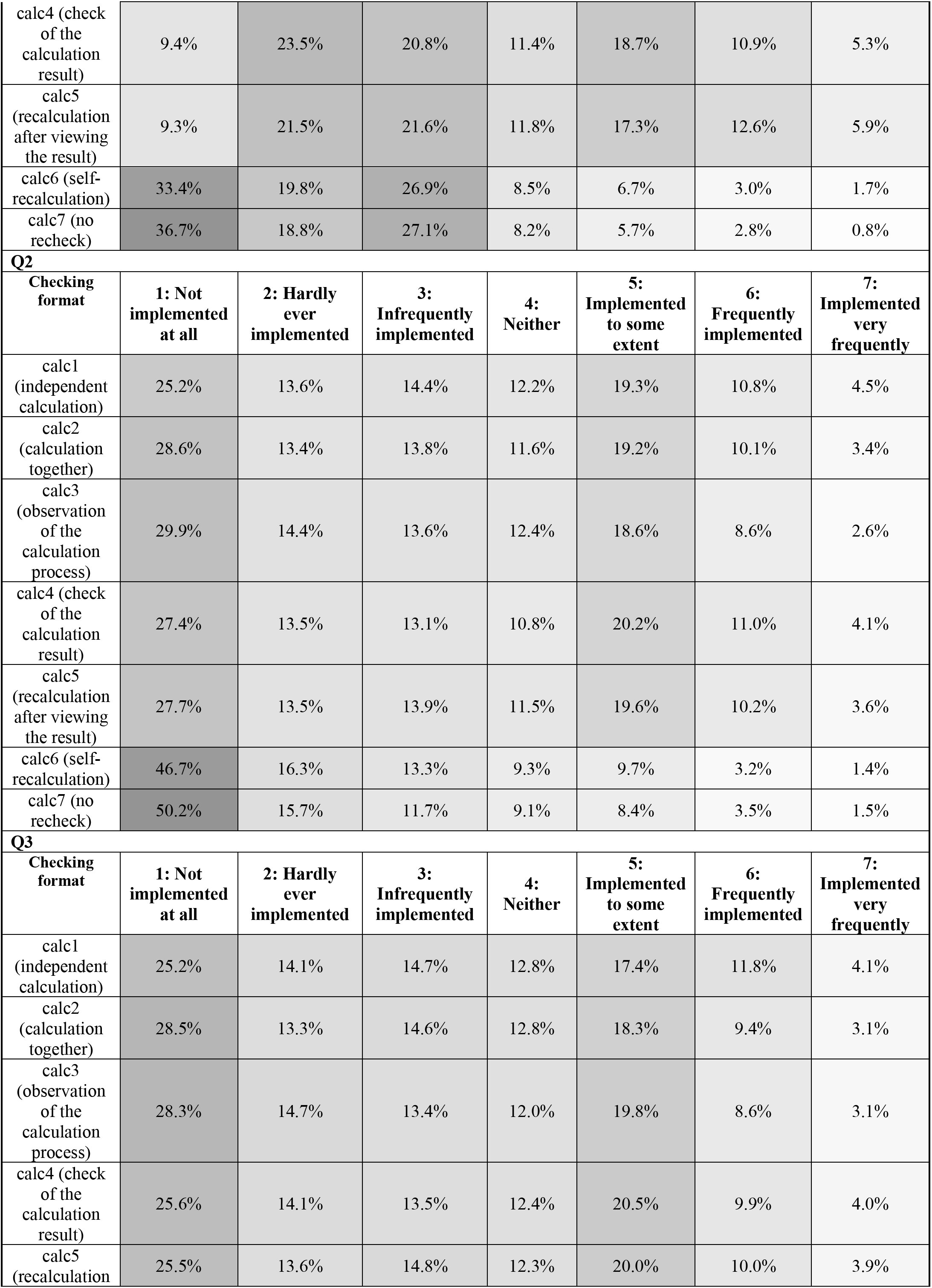
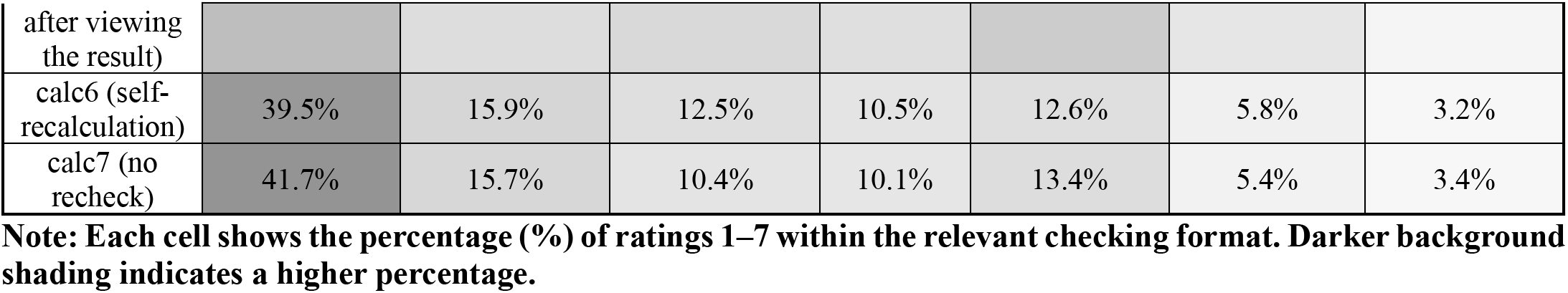
Rating distributions in the calculation stage.

##### (2) Preparation stage

For Q1, the proportions of ratings 5–7 were relatively high for prep1 and prep2, whereas the proportions of ratings 1–3 were high for prep3 and prep4. A similar distribution was observed for Q2. For Q3, the proportions of ratings 5–7 for prep3 and prep4 were higher than the corresponding proportions for Q1 and Q2, although the tendency for relatively higher ratings for prep1 and prep2 was maintained.

**Table D-4b.**
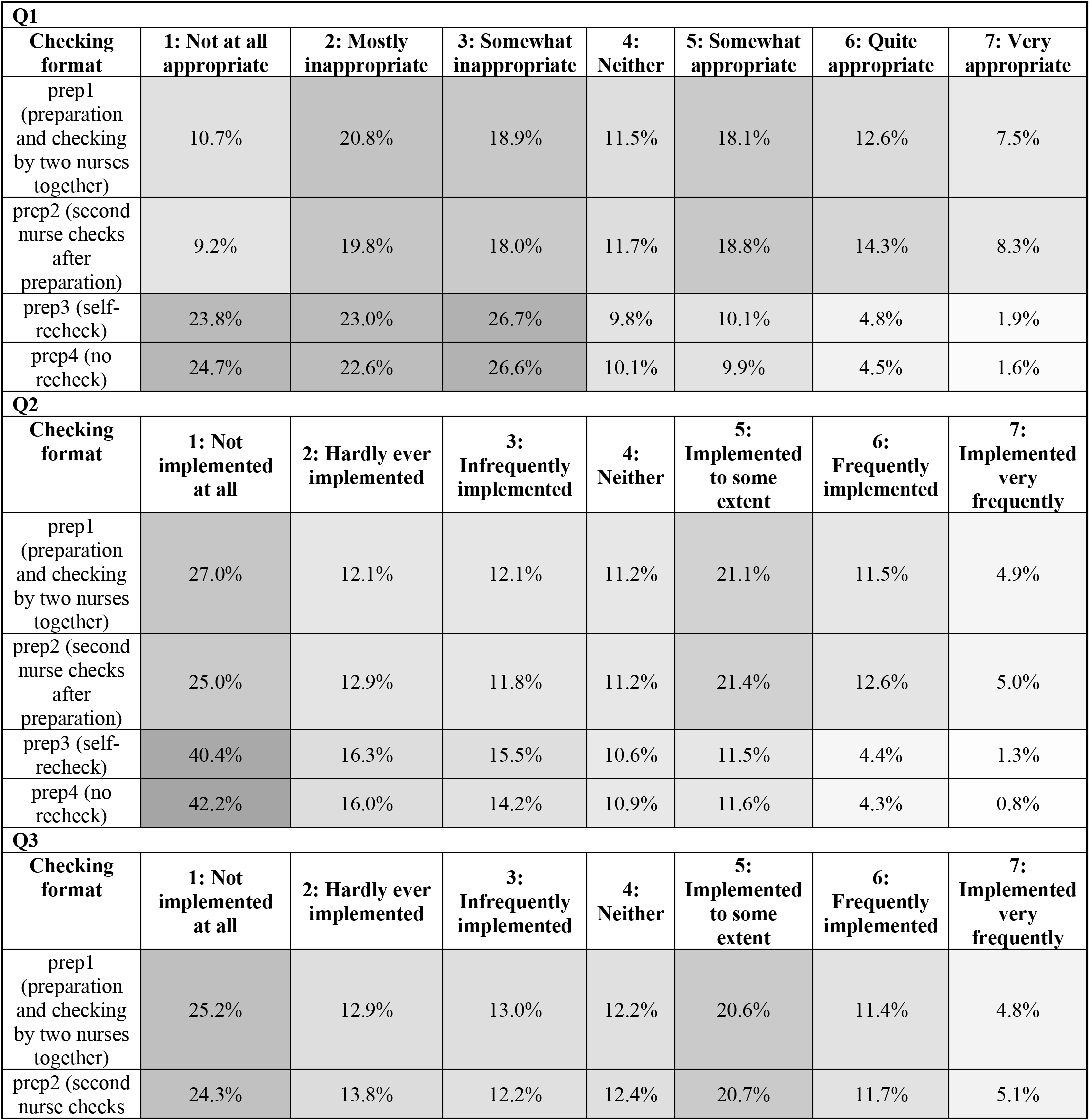
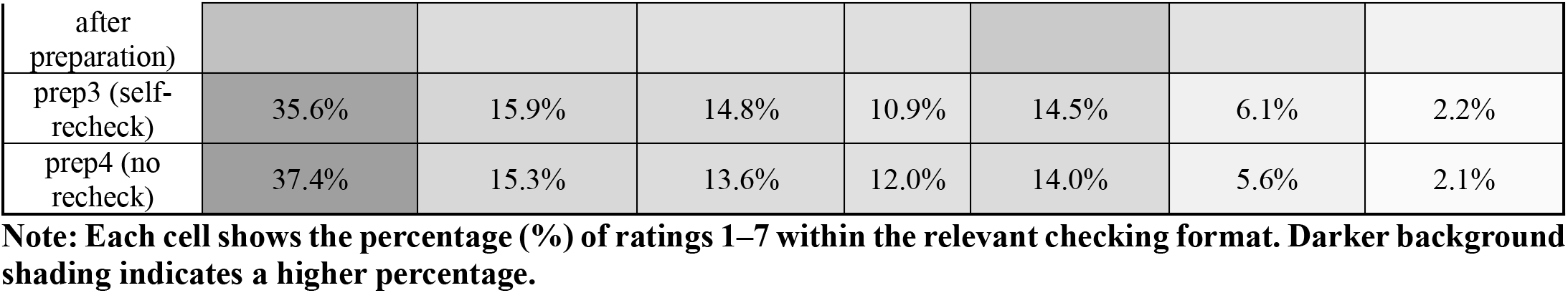
Rating distributions in the preparation stage.

##### (3) Administration stage

For Q1, the proportion of ratings 5–7 was higher for admin1 than for admin2 and admin3. For Q2, differences in the distributions among admin1–admin3 were smaller than those observed for Q1. For Q3, the proportions of ratings 5–7 were higher for admin2 and admin3 than for admin1.

**Table D-4c.** Rating distributions in the administration stage.

| **Q1** |  |  |  |  |  |  |  |
| --- | --- | --- | --- | --- | --- | --- | --- |
| Checking format | 1: Not at all appropriate | 2: Mostly inappropriate | 3: Somewhat inappropriate | 4: Neither | 5: Somewhat appropriate | 6: Quite appropriate | 7: Very appropriate |
| admin1 (pre-administration check by two nurses) | 11.8% | 19.7% | 18.6% | 10.5% | 15.4% | 13.4% | 10.7% |
| admin2 (self-recheck) | 19.1% | 22.0% | 24.7% | 11.1% | 13.8% | 7.2% | 2.0% |
| admin3 (no recheck) | 20.4% | 22.9% | 24.3% | 10.7% | 13.5% | 6.5% | 1.7% |
| **Q2** |  |  |  |  |  |  |  |
| Checking format | 1: Not implemented at all | 2: Hardly ever implemented | 3: Infrequently implemented | 4: Neither | 5: Implemented to some extent | 6: Frequently implemented | 7: Implemented very frequently |
| admin1 (pre-administration check by two nurses) | 30.7% | 14.0% | 13.3% | 11.0% | 16.8% | 9.7% | 4.5% |
| admin2 (self-recheck) | 35.0% | 14.3% | 13.3% | 11.1% | 16.4% | 7.5% | 2.5% |
| admin3 (no recheck) | 35.3% | 14.8% | 13.6% | 10.9% | 16.1% | 7.4% | 2.1% |
| **Q3** |  |  |  |  |  |  |  |
| Checking format | 1: Not implemented at all | 2: Hardly ever implemented | 3: Infrequently implemented | 4: Neither | 5: Implemented to some extent | 6: Frequently implemented | 7: Implemented very frequently |
| admin1 (pre-administration check by two nurses) | 32.3% | 15.9% | 15.1% | 11.7% | 15.1% | 6.8% | 3.1% |
| admin2 (self-recheck) | 29.2% | 13.7% | 12.6% | 12.1% | 19.0% | 9.7% | 3.7% |
| admin3 (no recheck) | 30.4% | 13.8% | 12.5% | 11.8% | 18.1% | 9.5% | 3.9% |

**Note:** Each cell shows the percentage (%) of ratings 1–7 within the relevant checking format. Darker background shading indicates a higher percentage.

### D.3 Estimated results and fit of the linear mixed-effects models

**Table D-5a.**
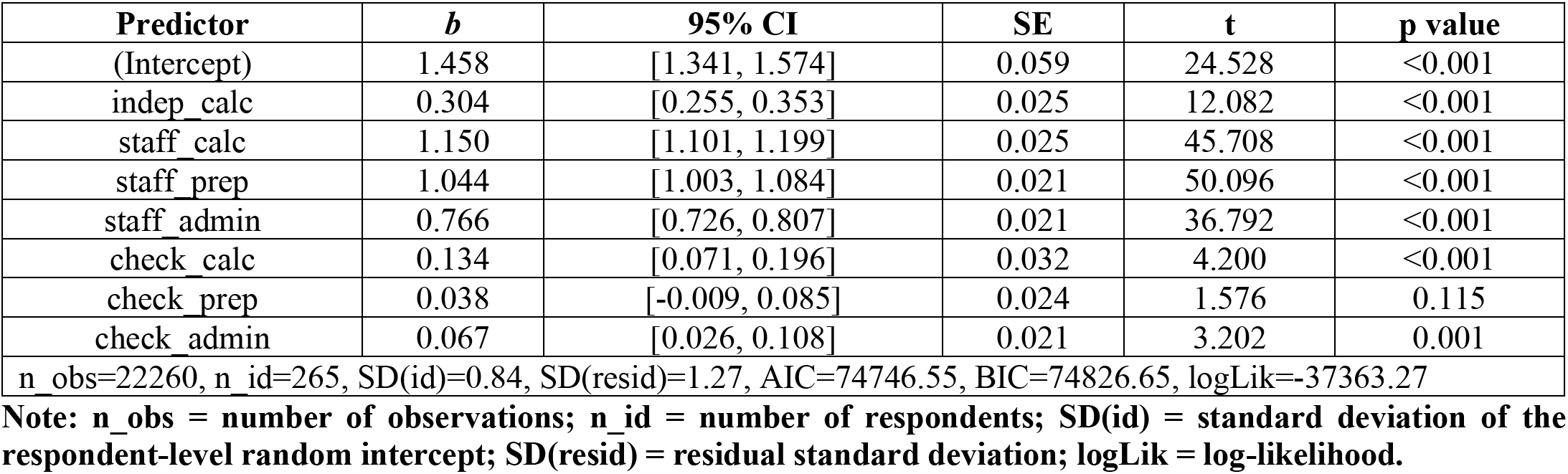
Model 1 results (Q1)

| Predictor | *b* | 95% CI | SE | *t* | p value |
| --- | --- | --- | --- | --- | --- |
| (Intercept) | 1.458 | [1.341, 1.574] | 0.059 | 24.528 | <0.001 |
| indep_calc | 0.304 | [0.255, 0.353] | 0.025 | 12.082 | <0.001 |
| staff_calc | 1.150 | [1.101, 1.199] | 0.025 | 45.708 | <0.001 |
| staff_prep | 1.044 | [1.003, 1.084] | 0.021 | 50.096 | <0.001 |
| staff_admin | 0.766 | [0.726, 0.807] | 0.021 | 36.792 | <0.001 |
| check_calc | 0.134 | [0.071, 0.196] | 0.032 | 4.200 | <0.001 |
| check_prep | 0.038 | [-0.009, 0.085] | 0.024 | 1.576 | 0.115 |
| check_admin | 0.067 | [0.026, 0.108] | 0.021 | 3.202 | 0.001 |
| n_obs=22260, n_id=265, SD(id)=0.84, SD(resid)=1.27, AIC=74746.55, BIC=74826.65, logLik=-37363.27 |  |  |  |  |  |

**Note:** n\_obs = number of observations; n\_id = number of respondents; SD(id) = standard deviation of the respondent-level random intercept; SD(resid) = residual standard deviation; logLik = log-likelihood.

**Table D-5b.** Model 2 results (Q1)

| Predictor | *b* | 95% CI | SE | *t* | p value |
| --- | --- | --- | --- | --- | --- |
| (Intercept) | 1.491 | [0.938, 2.044] | 0.281 | 5.308 | <0.001 |
| indep_calc | 0.304 | [0.255, 0.353] | 0.025 | 12.082 | <0.001 |
| staff_calc | 1.150 | [1.101, 1.199] | 0.025 | 45.708 | <0.001 |
| staff_prep | 1.044 | [1.003, 1.084] | 0.021 | 50.096 | <0.001 |
| staff_admin | 0.766 | [0.726, 0.807] | 0.021 | 36.792 | <0.001 |
| check_calc | 0.134 | [0.071, 0.196] | 0.032 | 4.200 | <0.001 |
| check_prep | 0.038 | [-0.009, 0.085] | 0.024 | 1.576 | 0.115 |
| check_admin | 0.067 | [0.026, 0.108] | 0.021 | 3.202 | 0.001 |
| beds_ord | -0.015 | [-0.073, 0.044] | 0.030 | -0.489 | 0.625 |
| beds_unk | 0.410 | [-0.954, 1.569] | 0.620 | 0.660 | 0.510 |
| tenure_ord | 0.053 | [-0.033, 0.139] | 0.044 | 1.220 | 0.224 |
| freq_reg_ord | -0.022 | [-0.153, 0.109] | 0.067 | -0.330 | 0.742 |
| freq_high_ord | -0.022 | [-0.131, 0.087] | 0.055 | -0.395 | 0.693 |
| n_obs=22260, n_id=265, SD(id)=0.84, SD(resid)=1.27, AIC=74753.33, BIC=74873.49, logLik=-37361.67 |  |  |  |  |  |

**Table D-5c.**
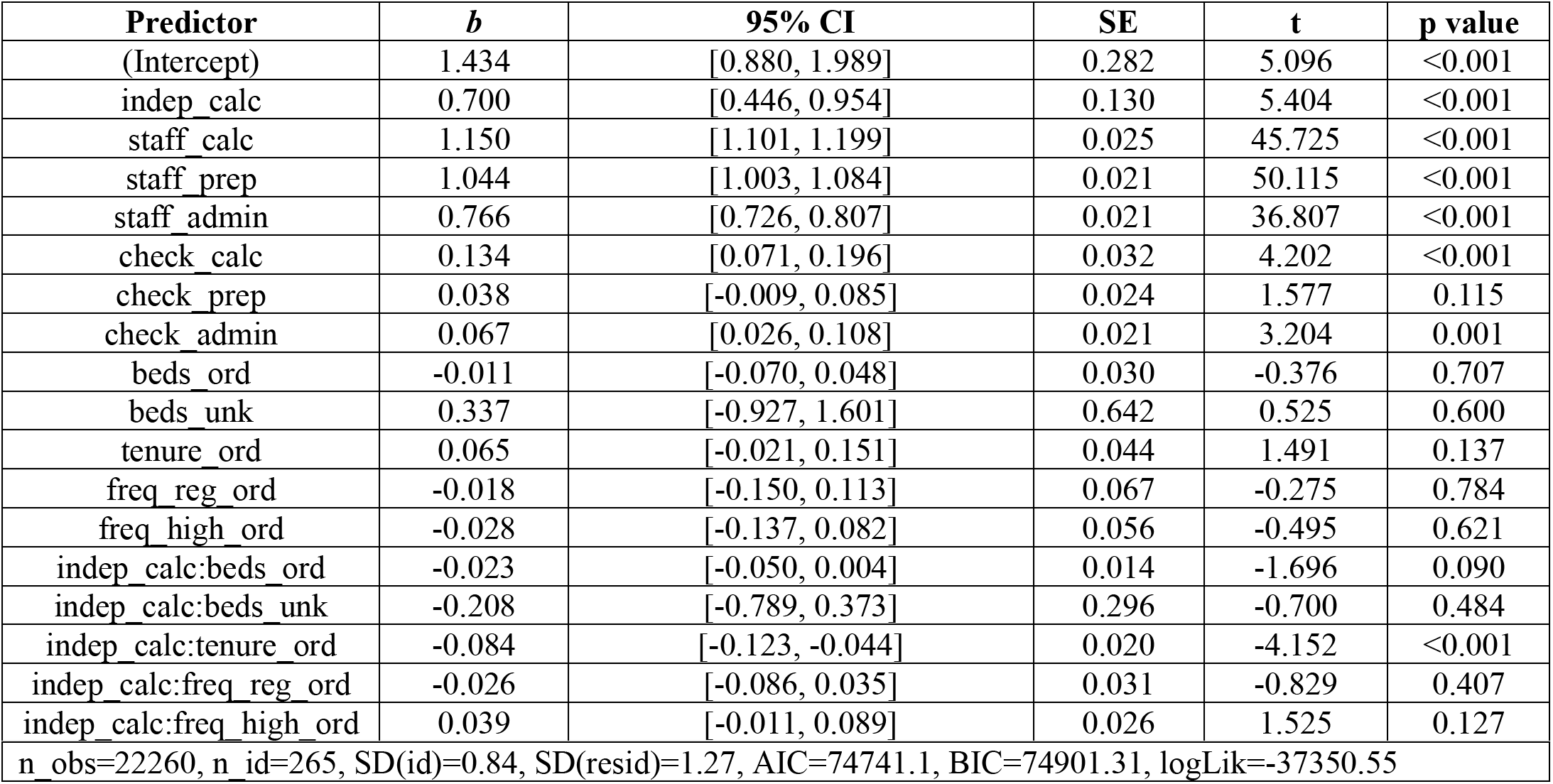
Model 3 results (Q1)

**Table D-6a.** Model 1 results (Q2)

| Predictor | *b* | 95% CI | SE | *t* | *p* value |
| --- | --- | --- | --- | --- | --- |
| (Intercept) | 1.667 | [1.514, 1.820] | 0.078 | 21.439 | <0.001 |
| indep_calc | 0.140 | [0.080, 0.200] | 0.031 | 4.563 | <0.001 |
| staff_calc | 0.889 | [0.828, 0.949] | 0.031 | 28.955 | <0.001 |
| staff_prep | 0.903 | [0.853, 0.953] | 0.025 | 35.531 | <0.001 |
| staff_admin | 0.244 | [0.194, 0.294] | 0.025 | 9.611 | <0.001 |
| check_calc | 0.081 | [0.004, 0.157] | 0.039 | 2.073 | 0.038 |
| check_prep | 0.053 | [-0.005, 0.110] | 0.029 | 1.800 | 0.072 |
| check_admin | 0.037 | [-0.013, 0.087] | 0.025 | 1.456 | 0.145 |

**Table D-6b.** Model 2 results (Q2)

| Predictor | *b* | 95% CI | SE | *t* | *p* value |
| --- | --- | --- | --- | --- | --- |
| (Intercept) | 0.981 | [0.161, 1.802] | 0.416 | 2.357 | 0.019 |
| indep_calc | 0.140 | [0.080, 0.200] | 0.031 | 4.563 | <0.001 |
| staff_calc | 0.889 | [0.828, 0.949] | 0.031 | 28.955 | <0.001 |
| staff_prep | 0.903 | [0.853, 0.953] | 0.025 | 35.531 | <0.001 |
| staff_admin | 0.244 | [0.194, 0.294] | 0.025 | 9.611 | <0.001 |
| check_calc | 0.081 | [0.004, 0.157] | 0.039 | 2.073 | 0.038 |
| check_prep | 0.053 | [-0.005, 0.110] | 0.029 | 1.800 | 0.072 |
| check_admin | 0.037 | [-0.013, 0.087] | 0.025 | 1.456 | 0.145 |
| beds_ord | -0.007 | [-0.089, 0.075] | 0.042 | -0.171 | 0.864 |
| tenure_ord | 0.142 | [0.031, 0.254] | 0.057 | 2.517 | 0.013 |
| freq_reg_ord | -0.074 | [-0.254, 0.106] | 0.091 | -0.807 | 0.421 |
| freq_high_ord | 0.163 | [0.019, 0.306] | 0.073 | 2.230 | 0.027 |

n\_obs=18637, n\_id=222, SD(id)=1.01, SD(resid)=1.42, AIC=66716.02, BIC=66825.68, logLik=-33344.01

**Table D-6c.** Model 3 results (Q2)

| Predictor | *b* | 95% CI | SE | *t* | *p* value |
| --- | --- | --- | --- | --- | --- |
| (Intercept) | 1.021 | [0.199, 1.843] | 0.417 | 2.448 | 0.015 |
| indep_calc | -0.137 | [-0.490, 0.215] | 0.180 | -0.764 | 0.445 |
| staff_calc | 0.889 | [0.829, 0.949] | 0.031 | 28.982 | <0.001 |
| staff_prep | 0.903 | [0.853, 0.953] | 0.025 | 35.561 | <0.001 |
| staff_admin | 0.244 | [0.194, 0.294] | 0.025 | 9.620 | <0.001 |
| check_calc | 0.081 | [0.004, 0.156] | 0.039 | 2.075 | 0.038 |
| check_prep | 0.053 | [-0.005, 0.110] | 0.029 | 1.803 | 0.071 |
| check_admin | 0.037 | [-0.013, 0.087] | 0.025 | 1.457 | 0.145 |
| beds_ord | -0.019 | [-0.101, 0.063] | 0.042 | -0.457 | 0.648 |
| tenure_ord | 0.155 | [0.043, 0.266] | 0.057 | 2.729 | 0.007 |
| freq_reg_ord | -0.083 | [-0.264, 0.097] | 0.092 | -0.908 | 0.365 |
| freq_high_ord | 0.168 | [0.024, 0.312] | 0.073 | 2.297 | 0.023 |
| indep_calc:beds_ord | 0.084 | [0.048, 0.119] | 0.018 | 4.634 | <0.001 |
| indep_calc:tenure_ord | -0.086 | [-0.134, -0.038] | 0.025 | -3.521 | <0.001 |
| indep_calc:freq_reg_ord | 0.066 | [-0.012, 0.144] | 0.040 | 1.668 | 0.095 |
| indep_calc:freq_high_ord | -0.037 | [-0.098, 0.025] | 0.032 | -1.155 | 0.248 |

n\_obs=18637, n\_id=222, SD(id)=1.01, SD(resid)=1.42, AIC=66685.72, BIC=66826.71, logLik=-33324.86

**Table D-7a.** Model 1 results (Q3)

| Predictor | *b* | 95% CI | SE | *t* | *p* value |
| --- | --- | --- | --- | --- | --- |
| (Intercept) | 2.378 | [2.237, 2.519] | 0.072 | 33.091 | <0.001 |
| indep_calc | 0.083 | [0.022, 0.143] | 0.031 | 2.669 | 0.008 |
| staff calc | 0.556 | [0.495, 0.616] | 0.031 | 17.959 | <0.001 |
| staff prep | 0.653 | [0.603, 0.704] | 0.026 | 25.504 | <0.001 |
| staff admin | -0.276 | [-0.326, -0.226] | 0.026 | -10.766 | <0.001 |
| check calc | 0.031 | [-0.045, 0.108] | 0.039 | 0.800 | 0.424 |
| check prep | 0.049 | [-0.009, 0.107] | 0.030 | 1.659 | 0.097 |
| check admin | 0.041 | [-0.010, 0.091] | 0.026 | 1.580 | 0.114 |
| n_obs=21405, n_id=255, SD(id)=0.99, SD(resid)=1.53, AIC=79879.46, BIC=79959.18, logLik=-39929.73 |  |  |  |  |  |

**Table D-7b.** Model 2 results (Q3)

| Predictor | *b* | 95% CI | SE | *t* | p value |
| --- | --- | --- | --- | --- | --- |
| (Intercept) | 1.922 | [1.255, 2.588] | 0.338 | 5.680 | <0.001 |
| indep calc | 0.083 | [0.022, 0.143] | 0.031 | 2.668 | 0.008 |
| staff calc | 0.556 | [0.495, 0.616] | 0.031 | 17.959 | <0.001 |
| staff prep | 0.653 | [0.603, 0.704] | 0.026 | 25.504 | <0.001 |
| staff admin | -0.276 | [-0.326, -0.226] | 0.026 | -10.766 | <0.001 |
| check calc | 0.031 | [-0.045, 0.108] | 0.039 | 0.800 | 0.424 |
| check prep | 0.049 | [-0.009, 0.107] | 0.030 | 1.659 | 0.097 |
| check admin | 0.041 | [-0.010, 0.091] | 0.026 | 1.580 | 0.114 |
| beds ord | 0.016 | [-0.055, 0.086] | 0.036 | 0.441 | 0.659 |
| tenure ord | 0.034 | [-0.067, 0.135] | 0.051 | 0.661 | 0.509 |
| freq reg ord | -0.012 | [-0.172, 0.147] | 0.081 | -0.153 | 0.878 |
| freq high ord | 0.104 | [-0.026, 0.234] | 0.066 | 1.582 | 0.115 |
| n_obs=21405, n_id=255, SD(id)=0.99, SD(resid)=1.53, AIC=79881.13, BIC=79992.73, logLik=-39926.56 |  |  |  |  |  |

**Table D-7c.**
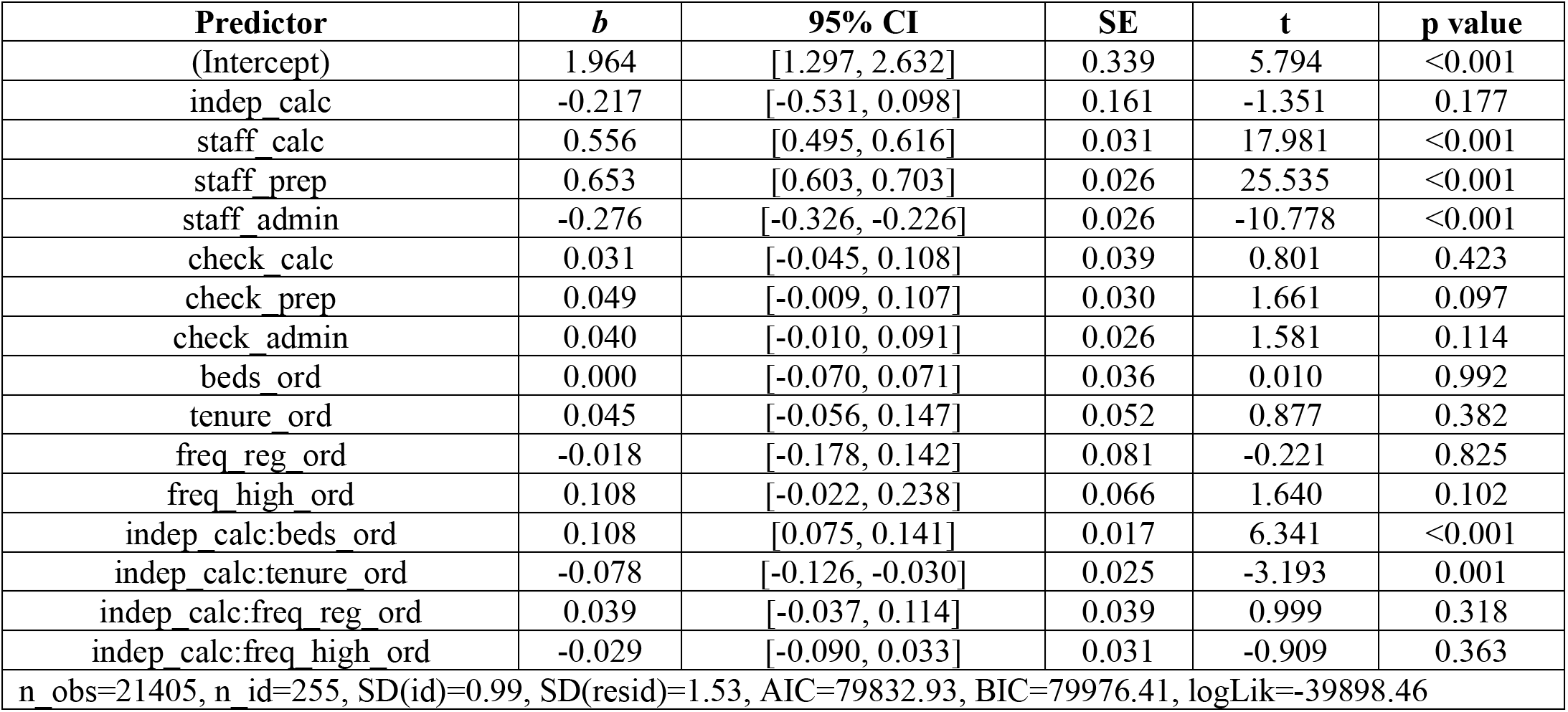
Model 3 results (Q3)

| Predictor | *b* | 95% CI | SE | *t* | p value |
| --- | --- | --- | --- | --- | --- |
| (Intercept) | 1.964 | [1.297, 2.632] | 0.339 | 5.794 | <0.001 |
| indep calc | -0.217 | [-0.531, 0.098] | 0.161 | -1.351 | 0.177 |
| staff calc | 0.556 | [0.495, 0.616] | 0.031 | 17.981 | <0.001 |
| staff prep | 0.653 | [0.603, 0.703] | 0.026 | 25.535 | <0.001 |
| staff admin | -0.276 | [-0.326, -0.226] | 0.026 | -10.778 | <0.001 |
| check calc | 0.031 | [-0.045, 0.108] | 0.039 | 0.801 | 0.423 |
| check prep | 0.049 | [-0.009, 0.107] | 0.030 | 1.661 | 0.097 |
| check admin | 0.040 | [-0.010, 0.091] | 0.026 | 1.581 | 0.114 |
| beds ord | 0.000 | [-0.070, 0.071] | 0.036 | 0.010 | 0.992 |
| tenure ord | 0.045 | [-0.056, 0.147] | 0.052 | 0.877 | 0.382 |
| freq reg ord | -0.018 | [-0.178, 0.142] | 0.081 | -0.221 | 0.825 |
| freq high ord | 0.108 | [-0.022, 0.238] | 0.066 | 1.640 | 0.102 |
| indep calc:beds ord | 0.108 | [0.075, 0.141] | 0.017 | 6.341 | <0.001 |
| indep calc:tenure ord | -0.078 | [-0.126, -0.030] | 0.025 | -3.193 | 0.001 |
| indep calc:freq reg ord | 0.039 | [-0.037, 0.114] | 0.039 | 0.999 | 0.318 |
| indep calc:freq high ord | -0.029 | [-0.090, 0.033] | 0.031 | -0.909 | 0.363 |
| n_obs=21405, n_id=255, SD(id)=0.99, SD(resid)=1.53, AIC=79832.93, BIC=79976.41, logLik=-39898.46 |  |  |  |  |  |

## Notes

### Competing Interest Statement

The authors have declared no competing interest.

### Author Declarations

Ethics committee of the Graduate School of Arts and Sciences, The University of Tokyo gave ethical approval for this work.

### Summary of Updates

ORCID iD was added for one of the authors.

